# Methamphetamine (MA) use, MA dependence, and MA-induced psychosis are associated with increasing aberrations in the compensatory immunoregulatory system and interleukin-1α and CCL5 levels

**DOI:** 10.1101/2023.03.26.23287766

**Authors:** Rasmon Kalayasiri, Kanokwan Dadwat, Supaksorn Thika, Sunee Sirivichayakul, Michael Maes

## Abstract

Comprehensive immunological profiles have not been studied in relation to methamphetamine (MA) use, MA dependency, or MA-induced psychosis (MAP). Using the BioPlex Pro Human Cytokine 48-Plex panel, this study measured M1 macrophage, T helper (Th)-1, Th-2, growth factor, and chemokine profiles, as well as the immune inflammatory response system (IRS) and compensatory immunoregulatory system (CIRS) in peripheral blood samples from patients with MA use (n=51), MA dependence (n=47), and MAP (n=43) in comparison with healthy controls (n=43). We discovered that persistent MA use had a robust dose-dependent suppressive impact on all immunological profiles, suggesting extensive immunosuppression. The most reliable biomarker profile of MA use is the combination of substantial CIRS suppression and a rise in selected pro-inflammatory cytokines, namely CCL27 (CTACK), CCL11 (eotaxin), and interleukin (IL)-1α. In addition, MA dependency is related with a more severe immunosuppression, as demonstrated by lower stem cell factor and higher IL-10 levels. MAP is related with a significant decrease in all immunological profiles, particularly CIRS, and an increase in CCL5 (RANTES), IL-1α, and IL-12p70 signaling. In conclusion, long-term MA use and dependency severely undermine immune homeostasis. This results in widespread immunosuppression, which may increase the likelihood of infectious and immune illness or exacerbate disorders such as hepatitis and AIDS. Elevated levels of CCL5, CCL11, CCL27, IL-1α, and/or IL-12p70 may be associated with severe peripheral (atherosclerosis, cutaneous inflammation, immune aberrations, hypospermatogenesis) and central (neuroinflammation, neurotoxic, neurodegenerative, depression, anxiety and psychosis) side effects. Our message: “cease using MA, or better yet, never begin using MA”.

## 1. Introduction

Methamphetamine (MA), a psychostimulant, is the second most frequently used illegal substance worldwide (Paulus and Stewart, 2020; Stoneberg et al., 2018). Between 2009 and 2018, the number of persons who used MA increased from 210 million to 265 million worldwide (UNODC, 2022). With at least 169 tonnes of MA seizures in 2020, seizures in East and Southeast Asia continued to rise (UNODC, 2021). Between 2016 and 2020, 72% of amphetamine-type stimulant seizures included MA, followed by 17% amphetamine and 4% ecstasy (UNODC, 2022). In Southeast Asia, particularly Thailand, the most popular drug has been MA speed tablets combining both MA and caffeine (Kalayasiri, 2016). In recent years, crystal meth (ice), a crystalline and pure version of MA, has gained popularity and has been introduced as a chemsex substance, including among those who inject drugs (Giorgetti et al., 2017; Werb et al., 2009).

An increase in dopamine, norepinephrine, and serotonin turnover causes the stimulant effects of MA on the central nervous system (Cruickshank and Dyer, 2009). In addition, the use of MA causes significant neurological and physical consequences, such as sleep disturbances, aggressive behavior, anxiety, sadness, depression, and psychosis, including delusions and hallucinations (Paulus and Stewart, 2020; Zeng et al., 2018). MA-induced psychosis (MAP) is characterized by persecuting delusions, hallucinations, and conceptual disorganization (Al-Hakeim et al., 2022; Chiang et al., 2019; Voce et al., 2019). Around 3– 23% of frequent users in a population sample of MA abusers reported MAP or psychotic symptoms (McKetin et al., 2006). Around 40% of persons with MA dependency in a hospital-based research group may have psychotic symptoms, such as paranoia, throughout their lifetime use of MA (Kalayasiri et al., 2014). MAP occurs over the course of MA use, including acute intoxication and chronic use (American Psychiatric Association, 2013). Typically, MAP may be present for a brief period, a few days to one month after the cessation of MA use; nevertheless, 38.8% of patients have a continuous course of psychosis, suggesting an association between MAP and primary psychotic disorders, such as schizophrenia (Kittirattanapaiboon et al., 2010).

A number of studies demonstrated that MA use may disrupt immunological homeostasis and suppress immune functions (Shi et al., 2022), including suppressing the production of T helper (Th)-1 cytokines (Yu et al., 2002), T-cell proliferation (Potula et al., 2018), and the number of lymphocytes and immune cells (Harms et al., 2012; In et al., 2005). Nitrostyrene, an amphetamine derivative, decreases interleukin (IL)-12 and IL-6 concentrations (Carter et al., 2002). Earlier research showed that MAP is associated with symptoms of immunological activation and inflammation, as well as neuroinflammation, as evidenced by elevated IL-6 and IL-8 (Yang et al., 2020), as well as elevated oxidative stress and decreased antioxidant defenses (Al-Hakeim et al., 2022). MA may affect antigen-presenting cells (APCs) in the brain, leading to an increase in pro-inflammatory cytokines such as IL-1β, IL-6, IL-8, interferons (IFN), and tumor necrosis factor (TNF)-α (Prakash et al., 2017).

MAP is often used and investigated as a model of schizophrenia because the overlapping symptoms and biomarker characteristics, such as elevated dopamine turnover (Ikeda et al., 2013). There is now evidence that schizophrenia, and particularly its more severe phenotype deficit schizophrenia, is a neuro-immune and neuro-oxidative disorder (Maes et al., 1994; Maes et al., 2022b; Roomruangwong et al., 2020), with elevated levels of pro-inflammatory cytokines such as IL-1β, IL-6, IL-12p70, and TNF-α (Monji et al., 2009; Song et al., 2009), activation of macrophage M1, Th-1, Th-2, Th-17, and T regulatory (Treg) phenotypes, as well as activation of the immune-inflammatory response system (IRS) and the compensatory immune-regulatory systems (CIRS), which tend to downregulate the IRS and prevent hyperinflammation (Roomruangwong et al., 2020). However, no research has studied the comprehensive immunological profiles of MA use, MA dependence, and MAP, including M1, Th-1, Th-2, Th-17, IRS, CIRS, chemokine, and growth factor profiles, to determine whether these conditions are characterized by immune activation or suppression. Hence, we explored peripheral blood immune profiles in MA use, MA dependence, and MAP as compared with healthy controls to examine whether these conditions are accompanied by immune activation or immunosuppression, and which immune profiles and cytokines are specifically involved in the three conditions.

## 2. Method

### 2.1 Participants

This research recruited 173 participants, 43 healthy controls and 141 Thai MA users from the Princess Mother National Institute on Drug Abuse Treatment (PMNIDAT), Thailand, including those with no MA dependency (n = 51), MA dependence (n = 47), and MAP (n = 43). A verbal notice was made at King Chulalongkorn Memorial Hospital to recruit healthy controls (HC) (n = 32) who had never taken MA in their lives and did not have any drug dependency except for a tobacco use disorder. By age and gender, the controls were matched with the MA group. All participants were between the ages of 18 and 65. Participants with severe neuropsychiatric disorders, such as schizophrenia, bipolar disorder, psycho-organic disorders, and schizo-affective disorder, as well as subjects with (auto)immune disorders, such as psoriasis, inflammatory-bowel disease, rheumatoid arthritis, and lupus erythematosus, were excluded from the study. We also omitted women who were pregnant or nursing. The data collection was between March - August 2022 and was approved by the Institutional Review Board of the Faculty of Medicine of Chulalongkorn University and the PMNIDAT (No. 28/2565).

### 2.2. Clinical assessment

Using the Thai version of the Semi-Structured Assessment of Drug Dependency and Alcoholism (SSADDA), demographics, drug use data including tobacco, alcohol, cannabis, and MA, and diagnoses were acquired (Kalayasiri et al., 2014; Malison et al., 2011). Based on the Fourth Version of the Diagnostic and Statistical Manual of Mental Disorders (DSM-IV), the SSADDA is a thorough interview to identify drug dependency and associated mental illnesses (American Psychiatric Association, 2000). Utilizing the SSADDA, patients meeting 6-7 criteria on the DSM-IV MA dependent scale were recruited for the MA dependence group, whereas those with 0-1 criteria were recruited for the no MA dependence group. Persons having a history of psychotic symptoms while taking MA, including hallucinations and/or delusions, were selected for the MAP group if they answered “yes” to any of the Psychotic Section of the SSADDA questions. Using the latter, we also determined the length of heavy MA use (in months), the daily quantity of MA (in milligrams; mg), and the forms of MA, such as crystal meth, speed pills, or both. We also evaluated lifetime and present alcohol and illicit substance use. We estimated the maximum number of alcoholic drinks per day; one standard alcoholic drink is equivalent to 10 grammes of pure alcohol. The interviewers were two psychologists with more than five years of training and experience doing SSADDA interviews. Before finishing the interview data, all interviews were cross-checked by several interviewers.

### 2.3 Measurement of cytokines

Early in the morning (between 8:00 and 9:00 a.m.), 5 ml of venous blood was drawn from each participant using a disposable syringe. Blood was stored at -80 °C until thawed for biomarker assays. The samples were placed at room temperature for 1 hour, then aliquots 25 μl. whole blood were diluted 1:4 (sample: diluent) after that the standard dilutions were prepared. The concentrations of 48 cytokines, chemokines and growth factors were measured using Bio-Plex Multiplex Immunoassays (Bio-Rad Laboratories Inc., USA). 50 μl of the diluted samples were combined with 50 μl of the microparticle cocktail (containing cytokines/chemokines per well of a 96-well plate) and incubated for 1 h at room temperature while shaking at 850 rpm The plates were rinsed three times, then 50 μl of diluted Streptavidin-PE was added to each well, and it was incubated for 10 minutes at room temperature. 125 μl of assay buffer was added, and the wells were shaken for 30 seconds at room temperature (850 rpm) prior to being read with the Bio-Plex® 200 System (Bio-Rad Laboratories, Inc.). In the present study, we employed the concentrations of the different analytes in the data analyses. Concentration that were not detectable (i.e., the lower limit out of range values) were censored and we imputed these censored data with the sensitivity values of each assay. Cytokines/chemokines/growth factors with concentration levels that were > 45% out of range were excluded from the analysis concerning the solitary analytes. Nevertheless, we also computed different immune profiles (see Introduction), including M1, Th-1, Th-2, IRS and CIRS profiles and all analytes (even those with < 45% detectable concentrations) may be used toward that end (Maes et al., 2022b; Thisayakorn et al., 2022), except those with <7% detectable concentrations (GM-CSF, IL-2, IL-3, IL-5, IL-15, and β-NGF). The reason is that even a smaller number of measurable concentrations (> more than 12) may contribute to the immune profiles. Electronic Supplementary File (ESF), Table 1 lists all cytokine/chemokines/growth factors measured here, as well as their names, gene IDs and % measurable analytes (> lower limit of out of range concentration). ESF, Table 2 shows how we constructed M1, Th-1, Th-2, IRS and CIRS immune profiles, based on our previous publications (Maes and Carvalho, 2018; Maes et al., 2022b; Thisayakorn et al., 2022) Unfortunately, we were unable to construct the Th-17 profile because its major players IL-6 and IL-17 were not measurable in many blood samples. We also computed the ratio z transformation of IL-12p70 – z IL-12p40, because IL-12 signaling via IL-12p70 may be inhibited via IL-12p40 (Zagozdzon and Lasek, 2016).

**Table 1.** Demographic and clinical data in healthy controls (HCs), patients with methamphetamine (MA)-use, MA-dependence, and MA-induced psychosis (MAP).

| Variables | HC<sup>A</sup><br>n=32 | MA use<sup>B</sup><br>n=51 | MA dependence<sup>C</sup><br>n=47 | MAP<sup>D</sup><br>n=43 | F/X<sup>2</sup> | df | p |
| --- | --- | --- | --- | --- | --- | --- | --- |
| **Demographics** |  |  |  |  |  |  |  |
| Female/male | 16/16 | 25/26 | 23/24 | 18/25 | 0.71 | 3 | 0.870 |
| Age (years) | 31.8 (10.7) | 33.6 (6.8)<sup>C</sup> | 29.0 (8.1)<sup>B</sup> | 31.0 (7.3) | 2.75 | 3/169 | 0.044 |
| Marital status (S/M/D) | 25/7/0 | 40/8/3 | 36/4/7 | 27/2/14 | FFHET | - | <0.001 |
| Body mass index (kilogram/metre<sup>2</sup>) | 26.2 (5.8) | 21.5 (3.8) | 22.4 (4.4) | 25.1 (4.9) | 9.40 | 3/169 | <0.001 |
| **MA use variables** |  |  |  |  |  |  |  |
| Duration of heaviest use (months) |  | 8.8 (30.3)<sup>C</sup> | 84.9 (87.9)<sup>B,D</sup> | 25.7 (47.1)<sup>C</sup> | 21.32 | 2/138 | <0.001 |
| Daily amount of heaviest use (milligrams) |  | 106.3 (113.9)<sup>C,D</sup> | 324.3 (343.7)<sup>B,D</sup> | 906.9 (908.0)<sup>B,C</sup> | 26.51 | 2/138 | <0.001 |
| DSM-IV criteria number<sup>a</sup> | 0.0<sup>B,C,D</sup> | 0.3 (0.5)<sup>A,C,D</sup> | 6.7 (0.7)<sup>A,B</sup> | 6.9 (0.4)<sup>A,B</sup> | KWT | - | <0.001 |
| MA dependence severity (z score) | -0.986 (0.0)<sup>C,D</sup> | -0.801 (0.270)<sup>C,D</sup> | 0.770 (0.688)<sup>A,B</sup> | 0.825 (0.780)<sup>A,B</sup> | KWT | - | <0.001 |
| Heaviest MA use (N/Cr/SP/Both) | 32/0/0/0 | 0/22/29/0 | 0/12/30/5 | 0/20/13/10 | - | - | - |
| **Alcohol use variables** |  |  |  |  |  |  |  |
| Maximum alcohol drinks per day<sup>b</sup> | 5.8 (6.7)<sup>B,C,D</sup> | 9.3 (13.6)<sup>A,D</sup> | 13.7 (12.2)<sup>A,D</sup> | 21.3 (24.9)<sup>A,B,C</sup> | 6.95 | 3/169 | <0.001 |
| Severity of alcohol use (z scores) | -0.336 (0.778)<sup>C</sup> | -0.243 (0.875)<sup>C</sup> | -0.007 (0.929)<sup>C</sup> | 0.546 (1.1541)<sup>A,B,C</sup> | 7.17 | 3/170 | <0.001 |
| Lifetime alcohol dependence (Y/N) | 0/31 | 2/46 | 8/379 | 21/21 | 40.93 | 3 | <0.001 |
| Current alcohol drinking (Y/N) | 2/29 | 2/46 | 1/44 | 0/43 | 2.92 | 3 | 0.404 |
| **Tobacco use variables** |  |  |  |  |  |  |  |
| Lifetime tobacco use (Y/N) | 8/24 | 43/8 | 43/4 | 41/1 | 69.62 | 3 | <0.001 |
| Current tobacco smoking (Y/N) | 8/24 | 15/36 | 12/35 | 4/39 | 6.00 | 3 | 0.112 |
| **Cannabis use variables** |  |  |  |  |  |  |  |
| Lifetime cannabis use (Y/N) | 7/25 | 18/33 | 36/11 | 32/11 | 37.32 | 3 | <0.001 |
| Current cannabis use (Y/N) | 0/32 | 0/51 | 2/45 | 0/43 | FFHET | - | 0.168 |

All data are shown as mean (SD), analyzed using analysis of variance (F) or Kruskal Wallis test (KWT); or as ratios, analyzed using $\chi^2$ tests or Fisher–Freeman–Halton exact test (FFHET); <sup>A,B,C,D</sup>: protected post-hoc comparisons among group means, S/M/D = single/married/divorced, N/Cr/SP/Both = none / crystal MA / speed pills / both, Y/N = yes/no; <sup>a</sup> Diagnostic and Statistical Manual on Mental Disorders (DSM-IV) symptom count or number of criteria for methamphetamine dependence; <sup>b</sup> alcohol standard drink or unit (1 drink = 10 gram of pure alcohol)

**Table 2.** Measurements of immune profiles in healthy controls (HCs), patients with MA-use, MA-dependence, and MA-induced psychosis (MAP).

| Z scores | HC<sup>A</sup><br>n=32 | MA use<sup>B</sup><br>n=51 | MA dependence<sup>C</sup><br>n=47 | MAP<sup>D</sup><br>n=43 | F | P-values |
| --- | --- | --- | --- | --- | --- | --- |
| M1 | 0.776 (0.146)<sup>B,C,D</sup> | 0.236 (0.117)<sup>A,D</sup> | 0.113 (0.118)<sup>A,D</sup> | -0.971 (0.125)<sup>A,B,C</sup> | 31.87 | <0.001** |
| Th-1 | 0.912 (0.139)<sup>B,C,D</sup> | 0.081 (0.111)<sup>A,D</sup> | 0.251 (0.112)<sup>A,D</sup> | -1.048 (0.118)<sup>A,B,C</sup> | 43.79 | <0.001** |
| IL-12<sub>70</sub> – IL-12<sub>40</sub> | -0.740 (0.155)<sup>B,C,D</sup> | -0.307 (0.124)<sup>A,C,D</sup> | 0.173 (0.125)<sup>A,B,D</sup> | 0.711 (0.132)<sup>A,B,C</sup> | 20.34 | <0.001** |
| Th-2 | 0.919 (0.144)<sup>B,C,D</sup> | 0.149 (0.115)<sup>A,D</sup> | 0.119 (0.116)<sup>A,D</sup> | -0.985 (0.123)<sup>A,B,C</sup> | 37.00 | <0.001** |
| IRS | 0.609 (0.159)<sup>B,D</sup> | 0.136 (0.127)<sup>A,D</sup> | 0.211 (0.128)<sup>D</sup> | -0.836 (0.135)<sup>A,B,C</sup> | 19.34 | <0.001** |
| CIRS | 1.010 (0.135)<sup>B,C,D</sup> | 0.085 (0.108)<sup>A,D</sup> | 0.181 (0.109)<sup>A,D</sup> | -1.042 (0.115)<sup>A,B,C</sup> | 48.65 | <0.001** |
| IRS/CIRS | -0.693 (0.175)<sup>B,C,D</sup> | 0.088 (0.140)<sup>A</sup> | 0.053 (0.141)<sup>A</sup> | 0.356 (0.149)<sup>A</sup> | 7.42 | <0.001** |
| Chemokines | 0.760 (0.141)<sup>B,C,D</sup> | 0.373 (0.113)<sup>A,C,D</sup> | 0.014 (0.114)<sup>A,B,D</sup> | -1.015 (0.120)<sup>A,B,C</sup> | 37.51 | <0.001** |
| Growth factors | 0.805 (0.144)<sup>C,D</sup> | 0.448 (0.115)<sup>C,D</sup> | -0.166 (0.116)<sup>A,B,D</sup> | -0.957 (0.122)<sup>A,B,C</sup> | 37.00 | <0.001** |
| PC_immune | 0.863 (0.140)<sup>B,C,D</sup> | 0.202 (0.112)<sup>A,D</sup> | 0.151 (0.113)<sup>A,D</sup> | -1.042 (0.119)<sup>A,B,C</sup> | 40.69 | <0.001** |

All results of univariate GLM analyses; shown are the estimated marginal mean (SE) values (in z scores) after covarying for body mass index, sex, age, current smoking, and current alcohol use (all df=3/164). <sup>A,B,C,D</sup>: protected post-hoc comparisons among group means.
M1: macrophage M1; Th: T helper, IL: interleukin; IRS: immune-inflammatory responses system; CIRS: compensatory immune-regulatory system; PC\_immune: first principal component extracted from the immune profiles.

### 2.4 Statistical analysis

Using Pearson’s product-moment correlation coefficients, correlations between variables were analyzed. The analysis of contingency tables was used to compare variables depending on their categories (Chi-square tests). Analysis of variance was employed to examine the differences in continuous variables across 4 study groups (HC, MA use, MA dependence, and MAP). Univariate generalized linear models taking into account the impacts of gender, age, BMI, current tobacco and alcohol use disorder, we examined the associations among these groups and cytokines and immune profiles. At a significance level of p 0.05, pairwise comparisons of group means were conducted to detect differences between the four study groups. Binary logistic regression analysis was performed to delineate the most important predictors of MAP versus MA without MAP (MA-MAP), and MA dependence versus MA use. Using the overfit criterion as entry and/or removal criterion (maximum effects number set at 5) we performed forward stepwise automatic linear modelling analyses (allowing for the confounders) and consequently, examined the model using a manual regression analyses. We checked the residual distributions of the final model, the variance inflation factor and tolerance in order to identify any collinearity or multicollinearity concerns, and heteroskedasticity using the White and modified Breusch-Pagan homoscedasticity tests. We calculated the partial regression analysis of immune data on clinical data, model statistics (F, df, and p values, total variance explained, which was used as effect size), and the standardized β coefficients, t statistics (with exact p-value) for each predictor. The significance threshold of all statistical analyses was calculated using two-tailed tests with a value of 0.05. Where needed, the cytokines/chemokines/growth factors were introduced in the statistical analyses after transformations, including log10, fractional rank-based normal transformations, and Winsorization. We used principal component analysis (PCA) as a feature reduction method and the first PC was considered to be adequate when the variance explained (VE) in the data was > 50%, and all variables have a loading > 0.66, the factoriability of the correlation matrix using the Kaiser-Meyer-Olkin (KMO) Measure of Sampling Adequacy, the Bartlett’s sphericity and the anti-image correlation matrix were accurate. All of the aforementioned statistical studies were conducted using IBM, SPSS windows version 28. An a priori sample size calculation was conducted using G*Power 3.1.9.4; for an analysis of covariance with 5 covariates, effect size of 0.27, alpha=0.05 (two tailed), power=0.8, and 4 groups the sample size should be at least 154.

## 3. Results

### Demographics and clinical data in the four study groups

**Table 1** shows the sociodemographic and clinical data of the 4 study groups. There were no significant differences in the sex ratio among groups, although there were some differences in age, BMI and marital status between the groups. MAP patients showed a significantly higher amount of daily MA use (in mg), alcohol drinks per day, and severity of alcohol use than patients without MAP. The severity of MA dependence and the DSM-IV number of dependence criteria were significantly higher in MA dependence and MAP than in those with MA use. There were significant differences in lifetime, but not current, tobacco and cannabis use among the four study groups.

### Immune profiles in MA use and dependence and MAP

**Table 2** shows the immune profiles of the four study groups. Using PCA, we were able to extract one PC from M1, Th-1, Th-2, IRS, CIRS, chemokine and growth factor profiles (VE=84.7%, all loadings > 0.878, KMO=0.896, bartlett’s test: χ2=1652.808, df=21, p<0.001), labeled PC_immune. All of the cytokine profiles including PC_immune, were significantly different between the study groups (p < 0.001). The MAP group had the lowest M1, Th-1, Th-2, IRS, CIRS, chemokine, growth factor, and PC_immune scores but higher IL12_70_ - IL20_40_ scores as compared with the three other groups. In addition, MA dependence and/or MA use had lower z-scores of M1, Th-1, Th-2, IRS, CIRS, chemokine, growth factor, and PC_immune scores than healthy controls. The PC_immune score was lower in MAP than in all other subjects. The IL12_70_ - IL20_40_ scores were higher in patients than controls. A post-hoc analysis to compute the achieved power for the least significant comparison in Table 2 (IRS/CIRS), showed that the achieved power (at α=0.05, 4 groups, 5 covariates, n=174) was 0.987. The achieved power for all other comparisons was 1.0. ESF, Figures S1-S36 show the levels of the measurable cytokines/chemokines/growth factors in controls, MA-use, MA dependence, and MAP.

### Intercorrelation matrix

**Table 3** shows the associations between MA use and dependence features and the immune profiles. The MA dose was significantly and negatively correlated with the M1, Th-1, Th-2, IRS, CIRS, chemokine, growth factor, and PC_immune profiles, and positively with IL12_70_ - IL20_40_ and the IRS/CIRS ratio. The duration of MA use showed an inverse association with the growth factor profile. The severity of MA dependence was significantly associated with M1, Th-1, Th-2, IRS, CIRS, chemokine, growth factor, and PC_immune profiles, and a positive correlation with IL12_70_ - IL20_40_ and the IRS/CIRS ratio.

**Table 3.** Intercorrelation matrix between methamphetamine (MA) use and dependence features and immune profiles

| Variables | MA amount of use <sup>a</sup> | MA duration of use <sup>b</sup> | MA dependence severity <sup>c</sup> |
| --- | --- | --- | --- |
| M1 | -0.338 (<0.001) | -0.083 (0.274) | -0.438 (<0.001) |
| Th-1 | -0.396 (<0.001) | -0.044 (0.560) | -0.402 (<0.001) |
| IL12<sub>70</sub> - IL12<sub>40</sub> | 0.321 (<0.001) | 0.138 (0.070) | 0.475 (<0.001) |
| Th-2 | -0.376 (<0.001) | -0.062 (0.419) | -0.430 (<0.001) |
| IRS | -0.280 (<0.001) | -0.046 (0.548) | -0.321 (<0.001) |
| CIRS | -0.378 (<0.001) | -0.089 (0.243) | -0.436 (<0.001) |
| IRS / CIRS | 0.169 (0.026) | 0.074 (0.330) | 0.199 (0.009) |
| Chemokines | -0.368 (<0.001) | -0.102 (0.182) | -0.508 (<0.001) |
| Growth factors | -0.382 (<0.001) | -0.175 (0.021) | -0.557 (<0.001) |
| PC_immune | -0.381 (<0.001) | -0.074 (0.331) | -0.449 (<0.001) |

Shown are Pearson's product moment correlation coefficients (with exact p value, all: n=174)
<sup>a</sup> Daily amount of heaviest use of MA; <sup>b</sup> Duration of heaviest use of MA; <sup>c</sup> z unit-weighted composite score based on daily amount of heaviest use of MA, duration of heaviest use of MA and the Diagnostic and Statistical Manual on Mental Disorders (DSM-IV) number of criteria for MA dependence
M1: macrophage M1; Th: T helper, IL: interleukin; IRS: immune-inflammatory responses system; CIRS: compensatory immune-regulatory system; PC\_immune: first principal component extracted from the immune profiles.

### Prediction of the general immune profile by MA and other clinical features

**Table 4** shows two different regression models predicting PC_immune. The first model was performed using MA features only and shows that 26.8% of the variance in PC_immune is explained by the regression on the number of dependency criteria and in addition by MA dose (both inversely). **Figure 1** shows the partial regression of PC_immune on MA dose. Regression #2 was performed with MA, alcohol and cannabis features as explanatory variables and shows that 40.1% of the variance in PC_immune is explained by the combined effects of alcohol dependence (positively) and an ordinal variable based on the 4 groups (negatively).

**Table 4.** Results of multiple regression analysis with the immune profile (PC_immune) as dependent variable

|  |  | Parameter estimates with statistics |  |  | Model statistics and effect size |  |  |  |
| --- | --- | --- | --- | --- | --- | --- | --- | --- |
| Dependent variable | Explanatory variables | $\beta$ | t | P-values | R<sup>2</sup> | F<sub>df</sub> | df | P-values |
| #1. PC_immune | Model |  |  |  | 0.268 | 31.11 | 2/170 | <0.001 |
|  | Number of criteria <sup>a</sup> | -0.189 | -2.03 | 0.044 | - | - |  | - |
|  | MA dose <sup>b</sup> | -0.366 | -3.93 | <0.001 | - | - |  | - |
| #2. PC_immune | Model |  |  |  | 0.401 | 54.56 | 2/163 | <0.001 |
|  | HC / MAU / MAD / MAP | -0.743 | -10.60 | <0.001 | - | - |  | - |
|  | Alcohol dependence | 0.198 | 2.94 | 0.004 | - | - |  | - |

PC\_immune: first principal component extracted from macrophage M1; Th-1 (T-helper), Th-2, immune-inflammatory responses system; compensatory immune-regulatory system, growth factors, and chemokines; MA = methamphetamine; <sup>a</sup> Diagnostic and Statistical Manual on Mental Disorders (DSM-IV) number of criteria for MA dependence; <sup>b</sup> Daily amount of heaviest use of MA; <sup>c</sup> Duration of heaviest use of MA; MA HC/use/dep/MIP: ordinal variable with HC: healthy control = 0, MAU: MA use = 1, MAD: MA dependence =2; and MAP: MA-induced psychosis =3.

**Figure 1.**
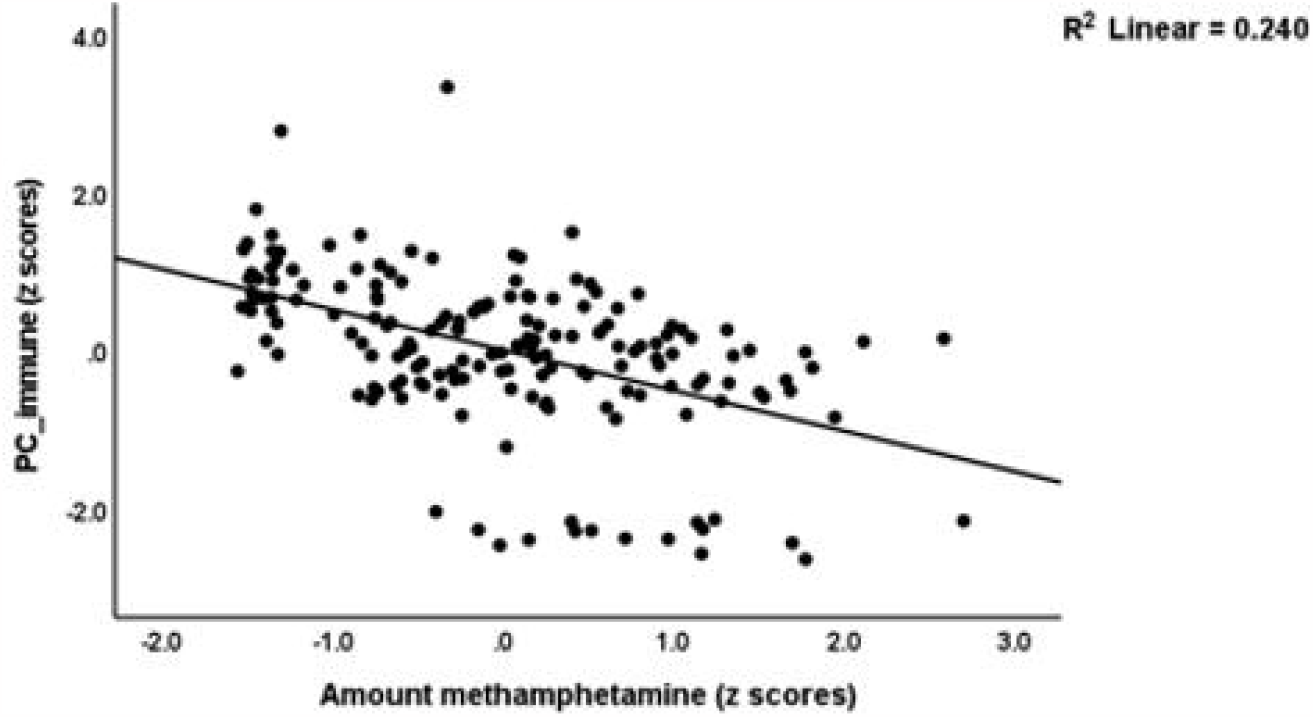
Partial regression of a generalized index of immune function (PC_immune) on the amount of methamphetamine

### Regression analyses with MAP and MA dependence as outcomes

**Table 5** shows the results of logistic binary regression analyses with MAP, MA dependence and MA use as dependent variables. Binary logistic regression #1 shows that MAP (reference group = MA patients without MAP) is predicted by lowered CIRS, and increased MA dose, number of DSM-IV MA dependence criteria and alcohol dependence (χ^2^=60.69, df=2, p<0.001, correctly classified or CC: 82.5%, Nagelkerke=0.492). Regression #2 shows that MAP was strongly predicted by Th-1 and CIRS profiles (inversely) and by CCL5 and IL-1α (positively) (χ^2^=96.41, df=4, p<0.001, correctly classified or CC: 87.3%, Nagelkerke=0.698). Regression #3 shows an alternative prediction of MAP with CIRS (inversely) and IL12_70_ - IL20_40_ and IL-1α (positively) as explanatory variables (χ^2^=103.4, df=3, p<0.001, correctly classified or CC: 86.7%, Nagelkerke=0.667). Regression #4 examined the prediction of MAP versus all other patients and controls and found that the same variables as shown in regression #2 predicted MAP (χ^2^=116.812, df=4, p<0.001, correctly classified or CC: 89.8%, Nagelkerke=0.726). Regression #5 shows that IRS/CIRS was the single best predictor of MAP (versus MA patients without MAP) (positive correlation). Binary logistic regression #6 examines the prediction of MA dependence (MA use as reference group) and shows that MA dependence was best predicted by SCF (inversely) and IL-10 (positively) (χ^2^=107.362, df=2, p<0.001, correctly classified or CC: 94.1%, Nagelkerke=0.888). The last regression (#7) shows that the best discrimination of MA patients versus healthy controls was obtained using CIRS (inversely) and CTACK+CCL11+IL-1α (positively) as predictors (χ^2^=92.793, df=2, p<0.001, correctly classified or CC: 93.1%, Nagelkerke=0.672).

**Table 5.** Results of logistic binary regression analysis with methamphetamine (MA)-induced psychosis (MAP), MA dependence, or MA use as dependent variables and immune profiles or single cytokines as explanatory variables.

| Dependent variable | ‡ Explanatory variables | B | SE | Wald<sub>df</sub> | df | p | OR | 95% CI |
| --- | --- | --- | --- | --- | --- | --- | --- | --- |
| #1. MAP versus MA-MAP | CIRS | -2.611 | 0.749 | 12.17 | 1 | <0.001 | 0.07 | 0.02; 0.32 |
|  | MA dose | 0.677 | 0.312 | 4.70 | 1 | 0.030 | 1.97 | 1.07; 3.63 |
|  | DSM-IV MA criteria | 2.803 | 1.348 | 4.33 | 1 | 0.038 | 16.50 | 1.18; 2.31 |
|  | Alcohol dependence | 0.340 | 0.146 | 5.46 | 1 | 0.019 | 1.41 | 1.06; 1.87 |
| #2. MAP vs MA-MAP | Th-1 | -2.904 | 0.990 | 8.60 | 1 | 0.003 | 0.06 | 0.01; 0.38 |
|  | CIRS | -2.100 | 1.008 | 4.34 | 1 | 0.037 | 0.12 | 0.02; 0.88 |
|  | CCL5 | 1.241 | 0.468 | 7.14 | 1 | 0.008 | 3.46 | 1.39; 8.60 |
| | IL-1α | 1.869 | 0.600 | 9.71 | 1 | 0.002 | 6.48 | 2.00; 20.99 |
| #3. MAP vs MA-MAP | IL-12<sub>70</sub>-IL12<sub>40</sub> | 0.549 | 0.279 | 3.91 | 1 | 0.047 | 1.73 | 1.01; 2.98 |
|  | CIRS | -3.330 | 0.633 | 27.66 | 1 | <0.001 | 0.036 | 0.01; 0.12 |
| | IL-1α | 1.784 | 0.533 | 11.23 | 1 | <0.001 | 5.96 | 2.10; 16.91 |
| #4. MAP vs MA-MAP + healthy controls | CIRS | -2.127 | 1.001 | 4.52 | 1 | <0.034 | 0.12 | 0.02; 0.85 |
|  | Th-1 | -2.901 | 0.991 | 8.57 | 1 | 0.003 | 0.06 | 0.008; 0.38 |
|  | CCL5 | 1.249 | 0.463 | 7.28 | 1 | 0.007 | 3.49 | 1.41; 8.65 |
| | IL-1α | 1.887 | 0.598 | 9.97 | 1 | 0.002 | 6.60 | 2.05; 21.29 |
| #5. MAP vs MA-MAP | IRS/CIRS | 0.501 | 0.228 | 4.81 | 1 | 0.028 | 1.65 | 1.05; 2.58 |
| #6. MA dependence vs MA use | SCF | -2.90 | 1.057 | 7.50 | 1 | 0.006 | 0.06 | 0.01; 0.44 |
|  | IL-10 | 8.70 | 2388 | 13.28 | 1 | <0.001 | 5998 | 5568; 64620 |
| #7. MA vs Healthy controls | CIRS | -4.121 | 0.730 | 31.85 | 1 | <0.001 | 0.02 | 0.004; 0.07 |
| | CCL27+CCL11+IL-1α | 2.679 | 0.705 | 14.44 | 1 | <0.001 | 14.58 | 3.66; 58.05 |

MAP versus MA-MAP: MAP versus MA use+MA dependence; M1: macrophage M1; Th: T helper, IL: interleukin; IRS: immune-inflammatory responses system; CIRS: compensatory immune-regulatory system; PC\_immune: first principal component extracted from the immune profiles; SCF:stem cell factor; CCL27 or CTACK: cutaneous T-cell attracting chemokine

## Discussion

### Immune profiles in MA use and dependence

The first major finding is that chronic MA use reduces considerably the first factor extracted from all immunological profiles (M1, Th-1, Th-2, IRS, CIRS, chemokine, and growth factors), indicating widespread immunosuppression. These findings expand upon those of recent papers demonstrating the many immunosuppressive effects of MA. MA at pharmacological doses exerts immunosuppressive effects on antigen-presenting cells, including macrophages and dendritic cells, reduces T cell proliferative activity, and inhibits receptor-mediated phagocytosis of antibody particles and MHC class II antigen presentation by T cells (Potula et al., 2018; Tallóczy et al., 2008). In addition, MA treatment of splenocytes has a substantial influence on antigen-induced proliferation and macrophage phagocytosis (Martinez et al., 2009). Two weeks of MA treatment inhibits lymphoproliferative responses to lipopolysaccharide (LPS) and concanavalin A in mice (In et al., 2005). In rodent models, injection of MA reduces the amount of T lymphocytes in the spleen, including CD4 and CD8 cells, and the number of macrophages, dendritic cells, and natural killer cells, with the latter exhibiting diminished reactivity (Harms et al., 2012). Furthermore, treatment of MA lowers the production of Th-1 cytokines, including IFN-γ, and IL-2, but had no effect on the production of IL-6 and IL-4 (Yu et al., 2002).

Importantly, immunosuppression in users of MA is strongly associated with increasing doses of MA (but not with duration of heaviest MA use) and with the severity of MA dependence. This indicates that MA has a strong dose-dependent suppressant effect on IRS and CIRS products. Overall, our data on M1 and Th-1 profiles are consistent with animal studies showing decreased M1 and Th-1 activity following MA exposure. Despite this, our findings indicate that chronic MA use additionally suppresses Th-2 and CIRS profiles, as well as the chemokine and growth factor subnetworks. This is important, as the latter subnetwork is intimately connected with the cytokine-chemokine network (Maes et al., 2022a). In addition, we discovered that prolonged MA use lowers the levels of several cytokines/chemokines/growth factors, such as FGF, GRO, IL-1β, sIL-1RA, sIL-2R, IL-4, IL-8, IL-12p40, IL-13, IL-18, LIF, CCL7 (MCP3), CSF, MIF, CXCL9 (MIG, SCF, and TRAIL.

Obviously, such a general decrease of immune processes may be accompanied by dysfunctions in host immunity, as observed in MA users, resulting in greater susceptibility to acquire new infections and a worsening of infectious diseases, including hepatitis and AIDS (Salamanca et al., 2015).

### Factors explaining MA-induced immunosuppression

Several variables may account for the significant immunosuppression generated by persistent MA use. First, MA may interfere with the cell cycle machinery, hence preventing T cell proliferation and a proper adaptive immune response (Potula et al., 2018). MA may reduce T cell cycle gene expression, including cyclin E, CDK2, and E2F, resulting in a prolonged G1/S phase transition (Potula et al., 2018). Second, injection of MA causes apoptosis in macrophages and T cells (Aslanyan et al., 2017; Potula et al., 2010). Third, MA-stimulated catecholamine turnover may suppress the production of IL-6, TNF-α, and sIL-1RA, therefore influencing the IRS and CIRS (Maes et al., 2000). Fourth, exposure to MA, a weak base, may interfere with the pH maintenance of the more acidic organelles, which govern protein breakdown and surface receptor expression (Martinez et al., 2009; Tallóczy et al., 2008). The effects of MA on the pH gradient may impair acidic organelles inside immune cells, MHC class II antigen processing, antigen presentation by dendritic cells to T cells, and ultimately the immunological response (Martinez et al., 2009; Tallóczy et al., 2008). Fifth, MA exposure may induce T cell mRNA expression of the trace amine-associated receptor 1 (TAAR1) (Sriram et al., 2016), which is involved in rheostasis, homeostasis, and cAMP signaling and interacts with monoamine turnover (Dodd et al., 2021; Maes et al., 2005). This indicates that MA-induced TAAR1 expression may downregulate Th-1-like cytokines such as IL-2 (Sriram et al., 2016).

### Signs of immune activation in MA use and dependence

Our second major observation is that persistent MA use is also associated with indicators of immunological activation. Various analytes were significantly elevated, such as CCL27 (CTACK) in MA users, and IL-1α, IL-12_p70_, and CCL5 (RANTES) in subjects with MA dependence. In addition, not all analytes were lowered in MA users, including CCL11 (eotaxin), IL-9, IL-16, CXCL10 (IP10), CCL3 (MIP1α), CCL4 (MIP1β), TNF-α and TNF-β (in both MA use and dependence), and G-CSF, HGF, CCL2 (MCP1), PDGF-BB, CXCL12 (SDF-1α), IL-9, and IL-10 (in MA dependence). These findings expand the findings of prior investigations. For example, treatment of MA to microglia markedly elevates IL-12_p70_ (Vargas et al., 2020), and MA exposure substantially increases IL-12 levels in the kidney and liver (but not spleen) of mice (Peerzada et al., 2013). Moreover, MA induces T cell proliferation in the brain via upregulating IL-15 in astrocytes (Bortell et al., 2017). Macaques afflicted with Simian Immunodeficiency Virus may produce more CCL5 if they are treated with MA (Najera et al., 2016). MA exposure elevates TNF-α levels in selected mouse brain areas, which are partially mediated by redox processes (Flora et al., 2002). Another research paper showed that MA exposure may considerably boost TNF-α production even in the presence of Th-1 suppression (Yu et al., 2002).

Considering that MA users have a generally suppressed immune system, the significant or relative increase in some cytokines/growth factors may be relevant. This is further supported by a higher IRS/CIRS ratio in MA users compared to healthy controls. Hence, despite the overall immunosuppression, there is a relative rise in pro-inflammatory cytokines/chemokines, as well as growth factors (G-CSF, PDGF-BB, and CXCL12 or SDF-1α) that may fuel the synthesis of these cytokines/chemokines. In fact, a combination of a decreased CIRS profile and an increase in pro-inflammatory cytokines/chemokines (CCL27 + CCL11 + IL-1α) is the best biomarker profile of MA use. It is noteworthy to note that CD4+ cells, which are inhibited during MA use, also exhibit indicators of activation, such as increased expression of CD150 and CD226 (Harms et al., 2012).

All in all, not only the generalized immunosuppression but also signs of immune activation are important to understand the pathophysiology of MA use. For example, CCL27 (CTACK or cutaneous T cell-attracting chemokine) is a cutaneous chemokine that attracts lymphocyte-associated antigen (CLA)+ memory cells, which are implicated in cutaneous inflammatory lesions (Morales et al., 1999). As such, increased CCL27 may perhaps play a role in MA-associated skin damage, including itching (due to meth mites), lesions, excoriations, and ulcers (Banyan Treatment Centers, 2021). IL-1α, IL-12_p70_, and CCL5 are pro-inflammatory cytokines/chemokines that may contribute to the inflammatory pathways and the accompanying brain dysfunctions (Salamanca et al., 2015) as well as peripheral inflammation-linked disorders, including cardiovascular diseases, which frequently occur in MA users (Al-Hakeim et al., 2022). CCL11 levels may be associated with greater anxiety, depression, and cognitive deficits among MA users (Huckans et al., 2015).

Moreover, the transition from moderate MA use to severe MA dependency (greater dosage and longer duration of MA use) is characterized by decreased SCF (stem cell factor) and increased IL-10 levels. SCF is a cytokine that plays a major role in hematopoiesis and spermatogenesis; consequently, decreased SCF may contribute to the decreased lymphoproliferative responses to MA (see discussion above) and the decreased sperm count, motility, and morphology in MA users (Allaeian Jahromi et al., 2022). In response to MA exposure, IL-10 levels are elevated in the plasma of both mice and humans (Loftis et al., 2011; Peerzada et al., 2013). Such a non-protective immunological response to IL-10 may restrict T cell proliferation even further.

### The immune profile of MAP

The third major finding of the study is that MAP is characterized by a) extremely reduced levels of M1, Th-1, Th-2, IRS, chemokine, and growth factor profiles, but especially the CIRS profile; b) an increase in a few selected cytokines/chemokines with systemic effects, including CCL5 and IL-1α, and IL-12p70 signaling; and c) an increased IRS/CIRS ratio. Alternatively stated, MA users with severe immunosuppression and increased levels of some pro-inflammatory cytokines are at risk for developing MAP. In addition, MAP is linked with increased MA use and severity of MA dependence, both of which are connected with rising immunosuppression and immune activation markers (e.g. IL-12 signaling).

IL-1α produced from peripheral blood may cross the BBB and get access to cortical brain cells (Banks et al., 1991). The production of IL-1α is mediated by elevated levels of damage-associated molecular patterns, necrosis, and necroptotic stimuli during sterile inflammation (Banks et al., 1993; Brough and Denes, 2015). Moreover, MA exposure may enhance the production of HMGB1, a significant DAMP, that contributes to neuroinflammation (Frank et al., 2016) and may promote necrosis/necroptosis via dopaminergic, oxidative stress, and AGE-RAGE pathways (Al-Hakeim et al., 2023; Davidson et al., 2001). Additionally, peripheral and central IL-1α elevations play a significant role in CNS inflammation, therefore contributing to acute and chronic brain diseases (Brough and Denes, 2015) Interestingly, IL-1α may generate CCL5, another cytokine related with MAP (Brough and Denes, 2015). As the levels of IL-1RA (which regulates IL-1-signaling; (Maes et al., 2012)) are significantly reduced in MAP, the effects of IL-1α and CCL5 may become more prominent. CCL5 (RANTES) is a chemokine that may cross the BBB and reach the brain parenchyma, and CCL5 and its receptor are expressed in glia, whilst microglia and astrocytes are capable of producing CCL5 (Bajetto et al., 2002; Dorf et al., 2000; Guo et al., 1998; Quaranta et al., 2023). Elevated CCL5 expression in the central nervous system is associated with increased neuroinflammation, cortical synaptic excitability, and hyperalgesia, and is implicated in neuroinflammatory and neurodegenerative disease (Bajetto et al., 2002; Benamar et al., 2008; Mori et al., 2016). In addition, enhanced IL-12 signaling contributes to neuroinflammation and neurodegeneration (Schneeberger et al., 2021; Turka et al., 1995; Vom Berg et al., 2012), and IL-12 (Turka et al., 1995) is, as CCL27, implicated in inflammatory skin lesions.

Despite the fact that it is now well-established that schizophrenia is an immunological condition and that first-episode schizophrenia and deficit schizophrenia are associated with IRS and CIRS activation (Maes, 2022; Maes et al., 1994; Noto et al., 2019), there are relatively few reports on the cytokine network in MAP. Yang et al. (2020) found that IL-6 and IL-8 were elevated in MAP patients and that sIL-2R was negatively correlated with positive symptoms (Yang et al., 2020). Psychosis in amphetamine-dependent women during early cessation is associated by increases in IL-10, TNF-α, and IL-5 (the latter cytokine was not measurable in our research) (Kuo et al., 2018). MAP is also accompanied by neuroinflammatory aberrations in the AAC-thalamus circuits (Burger et al., 2023).

## Conclusions

**Figure 2** summarizes our findings. Prolonged MA use disrupts immunological homeostasis, resulting in widespread immunosuppression (including elevated IL-10 levels and decreased SCF levels) and activation of specific cytokines/chemokines/growth factors (including CCL27, IL-1α, and CCL11). The increasing severity of MA dependence is accompanied with increased suppression of IRS and CIRS profiles and increases in IL-1α-CCL5 signaling and the IRS/CIRS ratio, which together appear to determine the onset of MAP. The findings show that despite the widespread immunosuppression, increased IL-1α-CCL5 signaling and IL-12p_70_ may contribute to MAP.

**Figure 2.**
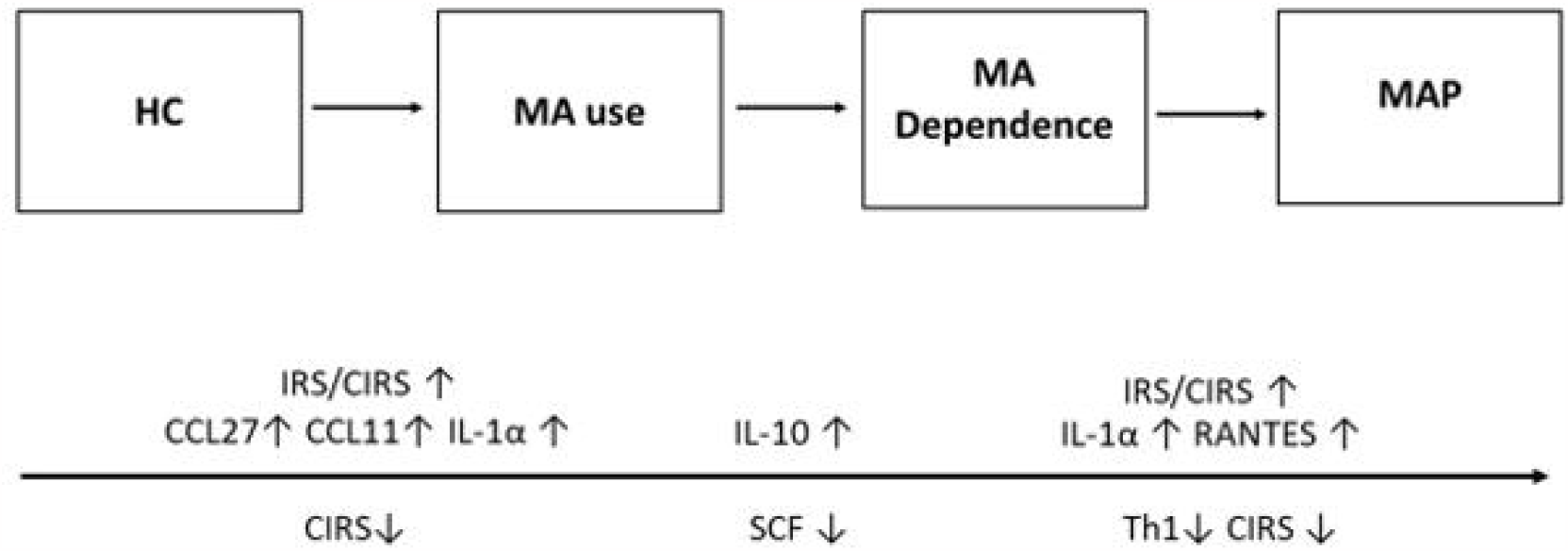
Summary of the findingsof the current study. HC: healthy controls; MA: methamphetamine; MAP: MA-induced psychosis IRS: immune-inflammatory responses system; CIRS: compensatory immunoregulatory system; IL: interleukin; Th: T helper: SCF: stem cell factor

## Supporting information

Electronic Supplementary File (ESF)

## Data Availability

All data produced in the present study are available upon reasonable request to the authors.

## Acknowledgment

The study is supported by the Centre for Addiction Studies, Department of Psychiatry, Faculty of Medicine, Chulalongkorn University and the Thai Health Promotion Foundation. We would like to thank Thitima Duangsanit and Somruk Kongchai for facilitate the process of data recruitment.

## Author Declarations

### Availability of data and materials

The dataset generated during and/or analyzed during the current study will be available from MM upon reasonable request and once the authors have fully exploited the dataset.

### Author’s Contributions

All authors contributed to the paper. MM and RK designed the study. RK supervised the recruitment of participants and sample collection. Participants were recruited and interviewed by ST and KD. Laboratory work was performed by SS. Statistical analyses were performed by M.M. All authors revised and approved the final draft.

## Compliance with Ethical Standards

### Research involving Human Participants and/or Animals

This study was approved by the Institutional Review Board (IRB) of Chulalongkorn University, Bangkok, Thailand and Princess Mother National Institute on Drug Abuse Treatment (No. 028/2565), which complies with the International Guideline for Human Research Protection as required by the Declaration of Helsinki.

### Informed consent

Before taking part in the study, all participants provided written informed consent.

## References

Al-Hakeim, H., Altufaili, M., Alhaideri, A., Almulla, A.F., Moustafa, S., Maes, M., 2023. Increased AGE-RAGE axis stress in methamphetamine (MA) abuse and MA-induced psychosis: associations with oxidative stress and increased atherogenicity. medRxiv, 2023-2001.

Al-Hakeim, H.K., Altufaili, M.F., Almulla, A.F., Moustafa, S.R., Maes, M., 2022. Increased lipid peroxidation and lowered antioxidant defenses predict methamphetamine induced psychosis. Cells 11, 3694.

Allaeian Jahromi, Z., Meshkibaf, M.H., Naghdi, M., Vahdati, A., Makoolati, Z., 2022. Methamphetamine Downregulates the Sperm-Specific Calcium Channels Involved in Sperm Motility in Rats. ACS omega 7, 5190-5196.

American Psychiatric Association, 2000. Diagnostic and Statistical Manual of Mental Disorders. 4th ed. American Psychiatric Association., Washington, D.C.

American Psychiatric Association, 2013. Diagnostic and Statistical Manual of Mental Disorders, Fifth Edition (DSM-5). American Psychiatric Publishing, Arlington, VA.

Aslanyan, L., Ekhar, V.V., DeLeon-Rodriguez, C.M., Martinez, L.R., 2017. Capsular specific IgM enhances complement-mediated phagocytosis and killing of Cryptococcus neoformans by methamphetamine-treated J774. 16 macrophage-like cells. International immunopharmacology 49, 77–84.

Bajetto, A., Bonavia, R., Barbero, S., Schettini, G., 2002. Characterization of chemokines and their receptors in the central nervous system: physiopathological implications. Journal of neurochemistry 82, 1311–1329.

Banks, W.A., Kastin, A.J., Gutierrez, E.G., 1993. Interleukin-1α in blood has direct access to cortical brain cells. Neuroscience letters 163, 41–44.

Banks, W.A., Ortiz, L., Plotkin, S., Kastin, A., 1991. Human interleukin (IL) 1 alpha, murine IL-1 alpha and murine IL-1 beta are transported from blood to brain in the mouse by a shared saturable mechanism. Journal of Pharmacology and Experimental Therapeutics 259, 988–996.

Banyan Treatment Centers, 2021. Taking care of your skin after meth addiction, Chicago.

Benamar, K., Geller, E.B., Adler, M.W., 2008. Elevated level of the proinflammatory chemokine, RANTES/CCL5, in the periaqueductal grey causes hyperalgesia in rats. European Journal of Pharmacology 592, 93–95.

Bortell, N., Basova, L., Semenova, S., Fox, H.S., Ravasi, T., Marcondes, M.C.G., 2017. Astrocyte-specific overexpressed gene signatures in response to methamphetamine exposure in vitro. Journal of neuroinflammation 14, 1–20.

Brough, D., Denes, A., 2015. Interleukin[1α and brain inflammation. IUBMB life 67, 323–330.

Burger, A., Lindner, M., Robertson, F., Blake, L., Williams, K., Naude, P., Temmingh, H., 2023. Comparison of schizophrenia and methamphetamine-induced psychosis: a proton magnetic resonance spectroscopy and cytokine study.

Carter, K., Finnon, Y., Daeid, N.N., Robson, D., Waddell, R., 2002. The effect of nitrostyrene on cell proliferation and macrophage immune responses. Immunopharmacology and immunotoxicology 24, 187–197.

Chiang, M., Lombardi, D., Du, J., Makrum, U., Sitthichai, R., Harrington, A., Shukair, N., Zhao, M., Fan, X., 2019. Methamphetamine[associated psychosis: Clinical presentation, biological basis, and treatment options. Human Psychopharmacology: Clinical and Experimental 34, e2710.

Cruickshank, C.C., Dyer, K.R., 2009. A review of the clinical pharmacology of methamphetamine. Addiction 104, 1085–1099.

Davidson, C., Gow, A.J., Lee, T.H., Ellinwood, E.H., 2001. Methamphetamine neurotoxicity: necrotic and apoptotic mechanisms and relevance to human abuse and treatment. Brain Research Reviews 36, 1–22.

Dodd, S., Carvalho, A.F., Puri, B.K., Maes, M., Bortolasci, C.C., Morris, G., Berk, M., 2021. Trace Amine-Associated Receptor 1 (TAAR1): A new drug target for psychiatry? Neuroscience & Biobehavioral Reviews 120, 537–541.

Dorf, M.E., Berman, M.A., Tanabe, S., Heesen, M., Luo, Y., 2000. Astrocytes express functional chemokine receptors. Journal of neuroimmunology 111, 109–121.

Flora, G., Lee, Y.W., Nath, A., Maragos, W., Hennig, B., Toborek, M., 2002. Methamphetamine-induced TNF-α gene expression and activation of AP-1 in discrete regions of mouse brain: potential role of reactive oxygen intermediates and lipid peroxidation. Neuromolecular medicine 2, 71–85.

Frank, M.G., Adhikary, S., Sobesky, J.L., Weber, M.D., Watkins, L.R., Maier, S.F., 2016. The danger-associated molecular pattern HMGB1 mediates the neuroinflammatory effects of methamphetamine. Brain, behavior, and immunity 51, 99–108.

Giorgetti, R., Tagliabracci, A., Schifano, F., Zaami, S., Marinelli, E., Busardò, F.P., 2017. When “chems” meet sex: a rising phenomenon called “chemsex”. Current neuropharmacology 15, 762–770.

Guo, H., Jin, Y.X., Ishikawa, M., Huang, Y.M., van der Meide, P.H., Link, H., Xiao, B.G., 1998. Regulation of beta-chemokine mRNA expression in adult rat astrocytes by lipopolysaccharide, proinflammatory and immunoregulatory cytokines. Scandinavian journal of immunology 48, 502–508.

Harms, R., Morsey, B., Boyer, C.W., Fox, H.S., Sarvetnick, N., 2012. Methamphetamine administration targets multiple immune subsets and induces phenotypic alterations suggestive of immunosuppression. PloS one 7, e49897.

Huckans, M., Fuller, B.E., Chalker, A.L., Adams, M., Loftis, J.M., 2015. Plasma inflammatory factors are associated with anxiety, depression, and cognitive problems in adults with and without methamphetamine dependence: an exploratory protein array study. Frontiers in psychiatry 6, 178.

Ikeda, M., Okahisa, Y., Aleksic, B., Won, M., Kondo, N., Naruse, N., Aoyama-Uehara, K., Sora, I., Iyo, M., Hashimoto, R., 2013. Evidence for shared genetic risk between methamphetamine-induced psychosis and schizophrenia. Neuropsychopharmacology 38, 1864–1870.

In, S.-W., Son, E.-W., Rhee, D.-K., Pyo, S., 2005. Methamphetamine Administration Produces Immunomodulation in Mice. Journal of Toxicology and Environmental Health, Part A 68, 2133–2145.

Kalayasiri, R., 2016. Addiction in Thailand, Neuropathology of drug addictions and substance misuse. London: Academic Press, pp. 1094–1100.

Kalayasiri, R., Verachai, V., Gelernter, J., Mutirangura, A., Malison, R.T., 2014. Clinical features of methamphetamine[induced paranoia and preliminary genetic association with DBH[1021 C→ T in a T hai treatment cohort. Addiction 109, 965–976.

Kittirattanapaiboon, P., Mahatnirunkul, S., Booncharoen, H., Thummawomg, P., Dumrongchai, U., Chutha, W., 2010. Long[term outcomes in methamphetamine psychosis patients after first hospitalisation. Drug and alcohol review 29, 456–461.

Kuo, S.C., Yeh, Y.W., Chen, C.Y., Huang, C.C., Ho, P.S., Liang, C.S., Lin, C.L., Yeh, T.C., Tsou, C.C., Yang, B.Z., Lu, R.B., Huang, S.Y., 2018. Differential effect of the DRD3 genotype on inflammatory cytokine responses during abstinence in amphetamine-dependent women. Psychoneuroendocrinology 97, 37–46.

Loftis, J.M., Choi, D., Hoffman, W., Huckans, M.S., 2011. Methamphetamine causes persistent immune dysregulation: a cross-species, translational report. Neurotoxicity research 20, 59–68.

Maes, M., 2022. Major neurocognitive psychosis: A novel schizophrenia endophenotype class that is based on machine learning and resembles Kraepelin’s and Bleuler’s conceptions. Acta Neuropsychiatrica, 1–15.

Maes, M., Carvalho, A.F., 2018. The compensatory immune-regulatory reflex system (CIRS) in depression and bipolar disorder. Molecular neurobiology 55, 8885–8903.

Maes, M., Kenis, G., Kubera, M., De Baets, M., Steinbusch, H., Bosmans, E., 2005. The negative immunoregulatory effects of fluoxetine in relation to the cAMP-dependent PKA pathway. International immunopharmacology 5, 609–618.

Maes, M., Lin, A., Kenis, G., Egyed, B., Bosmans, E., 2000. The effects of noradrenaline and alpha-2 adrenoceptor agents on the production of monocytic products. Psychiatry research 96, 245–253.

Maes, M., Meltzer, H., Bosmans, E., 1994. Immune[inflammatory markers in schizophrenia: comparison to normal controls and effects of clozapine. Acta Psychiatrica Scandinavica 89, 346–351.

Maes, M., Rachayon, M., Jirakran, K., Sodsai, P., Klinchanhom, S., Debnath, M., Basta-Kaim, A., Kubera, M., Almulla, A.F., Sughondhabirom, A., 2022a. Adverse childhood experiences predict the phenome of affective disorders and these effects are mediated by staging, neuroimmunotoxic and growth factor profiles. Cells 11, 1564.

Maes, M., Sirivichayakul, S., Matsumoto, A.K., Maes, A., Michelin, A.P., de Oliveira Semeão, L., de Lima Pedrão, J.V., Moreira, E.G., Barbosa, D.S., Geffard, M., 2022b. Correction to: Increased Levels of Plasma Tumor Necrosis Factor-α Mediate Schizophrenia Symptom Dimensions and Neurocognitive Impairments and Are Inversely Associated with Natural IgM Directed to Malondialdehyde and Paraoxonase 1 Activity. Molecular neurobiology 59, 1350.

Maes, M., Song, C., Yirmiya, R., 2012. Targeting IL-1 in depression. Expert opinion on therapeutic targets 16, 1097–1112.

Malison, R.T., Kalayasiri, R., Sanichwankul, K., Sughondhabirom, A., Mutirangura, A., Pittman, B., Gueorguieva, R., Kranzler, H.R., Gelernter, J., 2011. Inter-rater reliability and concurrent validity of DSM-IV opioid dependence in a Hmong isolate using the Thai version of the Semi-Structured Assessment for Drug Dependence and Alcoholism (SSADDA). Addictive behaviors 36, 156–160.

Martinez, L.R., Mihu, M.R., Gácser, A., Santambrogio, L., Nosanchuk, J.D., 2009. Methamphetamine enhances histoplasmosis by immunosuppression of the host. The Journal of infectious diseases 200, 131–141.

McKetin, R., McLaren, J., Lubman, D.I., Hides, L., 2006. The prevalence of psychotic symptoms among methamphetamine users. Addiction 101, 1473–1478.

Monji, A., Kato, T., Kanba, S., 2009. Cytokines and schizophrenia: Microglia hypothesis of schizophrenia. Psychiatry and clinical neurosciences 63, 257–265.

Morales, J., Homey, B., Vicari, A.P., Hudak, S., Oldham, E., Hedrick, J., Orozco, R., Copeland, N.G., Jenkins, N.A., McEvoy, L.M., 1999. CTACK, a skin-associated chemokine that preferentially attracts skin-homing memory T cells. Proceedings of the National Academy of Sciences 96, 14470–14475.

Mori, F., Nisticò, R., Nicoletti, C.G., Zagaglia, S., Mandolesi, G., Piccinin, S., Martino, G., Finardi, A., Rossini, P.M., Marfia, G.A., Furlan, R., Centonze, D., 2016. RANTES correlates with inflammatory activity and synaptic excitability in multiple sclerosis. Multiple Sclerosis Journal 22, 1405–1412.

Najera, J.A., Bustamante, E.A., Bortell, N., Morsey, B., Fox, H.S., Ravasi, T., Marcondes, M.C.G., 2016. Methamphetamine abuse affects gene expression in brain-derived microglia of SIV-infected macaques to enhance inflammation and promote virus targets. BMC immunology 17, 1–19.

Noto, M.N., Maes, M., Nunes, S.O.V., Ota, V.K., Rossaneis, A.C., Verri, W.A., Cordeiro, Q., Belangero, S.I., Gadelha, A., Bressan, R.A., Noto, C., 2019. Activation of the immune-inflammatory response system and the compensatory immune-regulatory system in antipsychotic naive first episode psychosis. European Neuropsychopharmacology 29, 416–431.

Paulus, M.P., Stewart, J.L., 2020. Neurobiology, clinical presentation, and treatment of methamphetamine use disorder: a review. JAMA psychiatry 77, 959–966.

Peerzada, H., Gandhi, J.A., Guimaraes, A.J., Nosanchuk, J.D., Martinez, L.R., 2013. Methamphetamine administration modifies leukocyte proliferation and cytokine production in murine tissues. Immunobiology 218, 1063–1068.

Potula, R., Haldar, B., Cenna, J.M., Sriram, U., Fan, S., 2018. Methamphetamine alters T cell cycle entry and progression: role in immune dysfunction. Cell death discovery 4, 44.

Potula, R., Hawkins, B.J., Cenna, J.M., Fan, S., Dykstra, H., Ramirez, S.H., Morsey, B., Brodie, M.R., Persidsky, Y., 2010. Methamphetamine causes mitrochondrial oxidative damage in human T lymphocytes leading to functional impairment. The journal of immunology 185, 2867–2876.

Prakash, M.D., Tangalakis, K., Antonipillai, J., Stojanovska, L., Nurgali, K., Apostolopoulos, V., 2017. Methamphetamine: effects on the brain, gut and immune system. Pharmacological research 120, 60–67.

Quaranta, D.V., Weaver, R.R., Baumann, K.K., Fujimoto, T., Williams, L.M., Kim, H.C., Logsdon, A.F., Omer, M., Reed, M.J., Banks, W.A., 2023. Transport of the Proinflammatory Chemokines CC Motif Chemokine Ligand 2 (MCP-1) and CC Motif Chemokine Ligand 5 (RANTES) across the Intact Mouse Blood-Brain Barrier Is Inhibited by Heparin and Eprodisate and Increased with Systemic Inflammation. Journal of Pharmacology and Experimental Therapeutics 384, 205–223.

Roomruangwong, C., Noto, C., Kanchanatawan, B., Anderson, G., Kubera, M., Carvalho, A.F., Maes, M., 2020. The role of aberrations in the immune-inflammatory response system (IRS) and the compensatory immune-regulatory reflex system (CIRS) in different phenotypes of schizophrenia: the IRS-CIRS theory of schizophrenia. Molecular neurobiology 57, 778–797.

Salamanca, S.A., Sorrentino, E.E., Nosanchuk, J.D., Martinez, L.R., 2015. Impact of methamphetamine on infection and immunity. Frontiers in neuroscience 8, 445.

Schneeberger, S., Kim, S., Eede, P., Boltengagen, A., Braeuning, C., Andreadou, M., Becher, B., Karaiskos, N., Kocks, C., Rajewsky, N., Heppner, F., 2021. The neuroinflammatory interleukin-12 signaling pathway drives Alzheimer’s disease-like pathology by perturbing oligodendrocyte survival and neuronal homeostasis.

Shi, S., Chen, T., Zhao, M., 2022. The Crosstalk Between Neurons and Glia in Methamphetamine-Induced Neuroinflammation. Neurochemical Research 47, 872–884.

Song, X.-Q., Lv, L.-X., Li, W.-Q., Hao, Y.-H., Zhao, J.-P., 2009. The interaction of nuclear factor-kappa B and cytokines is associated with schizophrenia. Biological psychiatry 65, 481–488.

Sriram, U., Cenna, J.M., Haldar, B., Fernandes, N.C., Razmpour, R., Fan, S., Ramirez, S.H., Potula, R., 2016. Methamphetamine induces trace amine[associated receptor 1 (TAAR1) expression in human T lymphocytes: role in immunomodulation. Journal of leukocyte biology 99, 213–223.

Stoneberg, D.M., Shukla, R.K., Magness, M.B., 2018. Global methamphetamine trends: an evolving problem. International Criminal Justice Review 28, 136–161.

Tallóczy, Z., Martinez, J., Joset, D., Ray, Y., Gácser, A., Toussi, S., Mizushima, N., Nosanchuk, J., Goldstein, H., Loike, J., 2008. Methamphetamine inhibits antigen processing, presentation, and phagocytosis. PLoS pathogens 4, e28.

Thisayakorn, P., Thipakorn, Y., Tantavisut, S., Sirivichayakul, S., Maes, M., 2022. Delirium due to hip fracture is associated with activated immune-inflammatory pathways and a reduction in negative immunoregulatory mechanisms. BMC psychiatry 22, 369.

Turka, L.A., Goodman, R.E., Rutkowski, J.L., Sima, A.A.F., Merry, A., Mitra, R.S., Wrone-Smith, T., Toews, G., Strieter, R.M., Nickoloff, B.J., 1995. Interleukin 12: A Potential Link between Nerve Cells and the Immune Response in Inflammatory Disorders. Molecular Medicine 1, 690–699.

UNODC, 2021. World Drug Report 2021 (Sales No. E.21.XI.8). Retrieved from United Nations publication.

UNODC, 2022. World Drug Report 2022 (Sales No. E.22.XI.8). Retrieved from United Nations publication.

Vargas, A.M., Rivera-Rodriguez, D.E., Martinez, L.R., 2020. Methamphetamine alters the TLR4 signaling pathway, NF-κB activation, and pro-inflammatory cytokine production in LPS-challenged NR-9460 microglia-like cells. Molecular immunology 121, 159–166.

Voce, A., Calabria, B., Burns, R., Castle, D., McKetin, R., 2019. A systematic review of the symptom profile and course of methamphetamine-associated psychosis: substance use and misuse. Substance use & misuse 54, 549–559.

Vom Berg, J., Prokop, S., Miller, K.R., Obst, J., Kälin, R.E., Lopategui-Cabezas, I., Wegner, A., Mair, F., Schipke, C.G., Peters, O., Winter, Y., Becher, B., Heppner, F.L., 2012. Inhibition of IL-12/IL-23 signaling reduces Alzheimer’s disease–like pathology and cognitive decline. Nature Medicine 18, 1812–1819.

Werb, D., Hayashi, K., Fairbairn, N., Kaplan, K., Suwannawong, P., Lai, C., Kerr, T., 2009. Drug use patterns among Thai illicit drug injectors amidst increased police presence. Substance abuse treatment, prevention, and policy 4, 1–5.

Yang, X., Zhao, H., Liu, X., Xie, Q., Zhou, X., Deng, Q., Wang, G., 2020. The Relationship Between Serum Cytokine Levels and the Degree of Psychosis and Cognitive Impairment in Patients With Methamphetamine-Associated Psychosis in Chinese Patients. Frontiers in Psychiatry 11.

Yu, Q., Zhang, D., Walston, M., Zhang, J., Liu, Y., Watson, R.R., 2002. Chronic methamphetamine exposure alters immune function in normal and retrovirus-infected mice. International immunopharmacology 2, 951–962.

Zagozdzon, R., Lasek, W., 2016. Biology of IL-12, in: Lasek, W., Zagozdzon, R. (Eds.), Interleukin 12: Antitumor Activity and Immunotherapeutic Potential in Oncology. Springer International Publishing, Cham, pp. 1–19.

Zeng, L., Tao, Y., Hou, W., Zong, L., Yu, L., 2018. Electro-acupuncture improves psychiatric symptoms, anxiety and depression in methamphetamine addicts during abstinence: a randomized controlled trial. Medicine 97.

