## Supplementary material for "Methamphetamine (MA) use, MA dependence, and MA-induced psychosis are associated with increasing aberrations in the compensatory immunoregulatory system and interleukin-1α and CCL5 levels": Electronic Supplementary File (ESF)

**ESF, Table 1**. Overview of the cytokines, chemokines and growth factors measured in the current study

| **Protein abbreviations** | **Gene Symbol** | **> OOR (%)** | **Protein name / alias** |
|---|---|---|---|
| **IFN-α2** | **IFNA2** | 42.0% | Interferon-α2 |
| **IFN-γ** | **IFNG** | 42.5% | Interferon-γ |
| **IL-1α** | **IL1A** | 100% | Interleukin-1α |
| **IL-1β** | **IL1B** | 99.4% | Interleukin-1β |
| **sIL-1RA** | **IL1RN** | 100% | Soluble interleukin-1 receptor antagonist |
| **IL-2** | **IL2** | <7% | Interleukin-2 |
| **IL-2RA** | **IL2RA** | 100% | Soluble interleukin-2 receptor |
| **IL-3** | **IL3** | 0% | Interleukin-3 |
| **IL-4** | **IL4** | 100% | Interleukin-4 |
| **IL-5** | **IL5** | <7% | Interleukin-5 |
| **IL-6** | **IL6** | 8% | Interleukin-6 |

| | | | |
|---|---|---|---|
| **IL-7** | **IL7** | 10.9% | Interleukin-7 |
| **IL-9** | **IL9** | 100% | Interleukin-9 |
| **IL-10** | **IL10** | 42% | Interleukin-10 |
| **IL-12p70** | **IL12RB1** | 47% | Interleukin-12 p70 |
| **IL-12p40** | **IL12RB1** | 57.5% | Interleukin-12 p40 |
| **IL-13** | **IL13** | 88.5% | Interleukin-13 |
| **IL-15** | **IL15** | <7% | Interleukin-15 |
| **IL-16** | **IL16** | 100% | Interleukin-16 |
| **IL-17** | **IL17A** | 8.6% | Interleukin-17 |
| **IL-18** | **IL18** | 100% | Interleukin-18 |
| **TNF-α** | **TNF** | 100% | Tumor necrosis factor-α |
| **TNF-β** | **LTA** | 100% | Tumor necrosis factor-β or lymphotoxin-alpha (LT-α) |
| **TRAIL** | **TNFSF10** | 100% | TNF-related apoptosis-inducing ligand (TRAIL) or tumor necrosis factor ligand superfamily member 10 (TNFSF10) |
| **LIF** | **LIF** | 100% | Interleukin inhibitory factor |
| **MIF** | **MIF** | 100% | Macrophage migration inhibitory factor-like protein (MIF) or glycosylation-inhibiting factor |

| | | | |
|---|---|---|---|
| **G-CSF** | **CSF3** | 100% | Granulocyte colony stimulating factor (G-CSF) or colony stimulating factor 3 (CSF3) |
| **M-CSF** | **CSF1** | 100% | Macrophage colony-stimulating factor (M-CSF) or colony stimulating factor 1 (CSF1) |
| **GM-CSF** | **CSF2** | <7% | Granulocyte-macrophage colony-stimulating factor (GM-CSF) or colony-stimulating factor 2 (CSF2) |
| **CCL2 or MCP1** | **CCL2** | 100% | C-C motif chemokine ligand 2 (CCL2) or monocyte chemoattractant protein 1 (MCP1) |
| **CCL3 or MIP-1α** | **CCL3** | 99.4% | C-C motif Chemokine ligand 3 (CCL3) or macrophage inflammatory protein 1-alpha (MIP-1α) |
| **CCL4 or MIP-1β** | **CCL4** | 100% | C-C motif chemokine ligand 4 (CCL4) or macrophage inflammatory protein 1β (MIP-1β) or lymphocyte activation gene 1 protein |
| **CCL5 or RANTES** | **CCL5** | 71.8% | C-C motif chemokine ligand 5 (CCL5) or regulated upon activation, normally T-expressed, and presumably Secreted (RANTES) |
| **CCL7 or MCP3** | **CCL7** | 78.2% | C-C motif chemokine ligand 7 (CCL7) or monocyte-chemotactic protein 3 (MCP3). |
| **CCL11 or Eotaxin** | **CCL11** | 100% | C-C motif chemokine ligand 11 (CCL11) or eosinophil chemotactic protein |
| **CCL27 or CTACK** | **CCL27** | 100% | C-C motif chemokine ligand 27 (CCL27) or cutaneous T-cell attracting chemokine (CTACK) |
| **CXCL1 or GRO-α** | **CXCL1** | 83.9% | C-X-C motif chemokine 1 (CXCL1) or growth-regulated alpha protein (GRO) |
| **CXCL8 or IL-8** | **CXCL8** | 99.4% | C-X-C motif chemokine ligand 8 (CXCL8) or interleukin-8 (IL-8) |
| **CXCL9 or MIG** | **CXCL9** | 100% | C-X-C motif chemokine ligand 9 (CXCL9) or monokine induced by gamma interferon (MIG) |
| **CXCL10 or IP10** | **CXCL10** | 100% | C-X-C motif chemokine ligand 10 (CXCL10) or Interferon gamma-induced protein 10 (IP10) |
| **CXCL12 or SDF-1α** | **CXCL12** | 100% | C-X-C motif chemokine 12 (CXCL12) or stromal cell-derived factor 1 (SDF-1α) |

| | | | |
|---|---|---|---|
| **FGF** | **FGF2** | 100% | Fibroblast growth factor 2 (FGF) or basic fibroblast growth factor |
| **HGF or SF** | **HGF** | 100% | Hepatocyte growth factor (HGF) or scatter factor (SF) |
| **NGF** | **NGF** | <7% | β-nerve growth factor (NGF) |
| **PDGF** | **PDGFA** | 99.4% | Platelet derived growth factor (PDGF) |
| **SCF** | **KITLG** | 100% | Stem cell factor (SCF) or Kit ligand (KITLG) |
| **SCGF-β or CLEC11A** | **CLEC11A** | 100% | Stem cell growth factor (SCGF) or C-type lectin domain family 11 member A (CLEC11A) |
| **VEGF** | **VEGFA** | 15.5% | Vascular endothelial growth factor (VEGF) |

**ESF, Table 2**. Composite scores used in the present study

| Immune Profiles | Members |
|---|---|
| **M1 macrophage** | IL-1α, IL-1β, sIL-1RA, IL-6, TNF-α, CXCL8, CXCL10, CCL2, CCL3, CCL4 |
| **T helper-1** | IL-2, sIL-2R, IFN-α, IFN-γ, IL-12p70, IL12p40, IL-16 |
| **T helper-2** | IL-4, IL-9, IL-13 |
| **IRS** | IL-1α, IL-1β, IL-6, LIF, IL-7, TNF-α, TNF-β, TRAIL, IFN-α2, IFN-γ, IL-12p70, IL-16, IL-18, G-CSF, M-CSF, CCL2, CCL3, CCL4, CCL5, CCL7, CC11, CCL, 27, CXCL1, CXCL8, CXCL9, CXCL10, CXCL12 |
| **CIRS** | IL-4, IL-9, IL-10, sIL-1RA, IL-12p40, sIL-2R |
| **Chemokines** | All CCL and all CXCL members |
| **Growth factors** | FGF, HGF, PDGF, SCF, SCGF, VEGF |

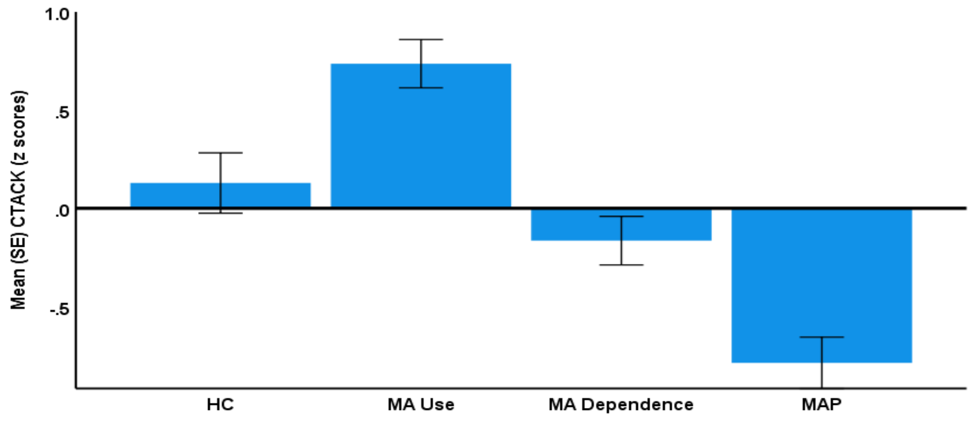


Figure S1. CCL27 or CTACK levels in healthy controls (HC) and people with methamphetamine (MA) use, MA dependence, and MA-induced psychosis (MAP) (F=23.87, df=3/164, p<0.001).

**Pairwise Comparisons**

Dependent Variable: CTACK

| (I) HC_Abuse_Dependence_Psychosis | (J) HC_Abuse_Dependence_Psychosis | Mean Difference (I-J) | Std. Error | Sig.[b] | 95% Confidence Interval for Difference[b] Lower Bound | Upper Bound |
|---|---|---|---|---|---|---|
| 0 | 1 | -.608* | 0.202 | 0.003 | -1.006 | -0.209 |
| | 2 | 0.293 | 0.200 | 0.145 | -0.102 | 0.688 |
| | 3 | .915* | 0.199 | <.001 | 0.523 | 1.308 |
| 1 | 0 | .608* | 0.202 | 0.003 | 0.209 | 1.006 |
| | 2 | .901* | 0.174 | <.001 | 0.557 | 1.245 |
| | 3 | 1.523* | 0.185 | <.001 | 1.158 | 1.888 |
| 2 | 0 | -0.293 | 0.200 | 0.145 | -0.688 | 0.102 |
| | 1 | -.901* | 0.174 | <.001 | -1.245 | -0.557 |
| | 3 | .622* | 0.181 | <.001 | 0.265 | 0.980 |
| 3 | 0 | -.915* | 0.199 | <.001 | -1.308 | -0.523 |
| | 1 | -1.523* | 0.185 | <.001 | -1.888 | -1.158 |
| | 2 | -.622* | 0.181 | <.001 | -0.980 | -0.265 |

Based on estimated marginal means

*. The mean difference is significant at the 0.05 level.

b. Adjustment for multiple comparisons: Least Significant Difference (equivalent to no adjustments).

HC=0, MA use=1, MA dependence=2, MAP=3

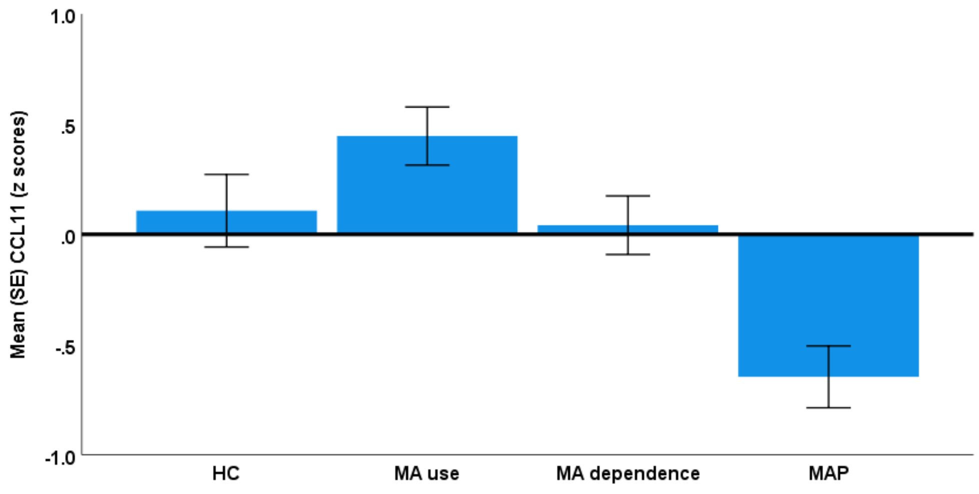


Figure S2. CCL11 levels in healthy controls (HC) and people with methamphetamine (MA) use, MA dependence, and MA-induced psychosis (MAP) (F=10.64, df=3/164, p<0.001).

**Pairwise Comparisons**

Dependent Variable: CCL11

| (I) HC_Abuse_Dependence_Psychosis | (J) HC_Abuse_Dependence_Psychosis | Mean Difference (I-J) | Std. Error | Sig.[b] | 95% Confidence Interval for Difference[b] | |
|---|---|---|---|---|---|---|
| | | | | | Lower Bound | Upper Bound |
| 0 | 1 | -0.339 | 0.217 | 0.120 | -0.767 | 0.089 |
| | 2 | 0.066 | 0.215 | 0.760 | -0.358 | 0.489 |
| | 3 | .754* | 0.213 | 0.001 | 0.333 | 1.175 |
| 1 | 0 | 0.339 | 0.217 | 0.120 | -0.089 | 0.767 |
| | 2 | .404* | 0.187 | 0.032 | 0.036 | 0.773 |
| | 3 | 1.092* | 0.198 | 0.000 | 0.701 | 1.484 |
| 2 | 0 | -0.066 | 0.215 | 0.760 | -0.489 | 0.358 |
| | 1 | -.404* | 0.187 | 0.032 | -0.773 | -0.036 |
| | 3 | .688* | 0.194 | 0.001 | 0.305 | 1.071 |
| 3 | 0 | -.754* | 0.213 | 0.001 | -1.175 | -0.333 |
| | 1 | -1.092* | 0.198 | 0.000 | -1.484 | -0.701 |
| | 2 | -.688* | 0.194 | 0.001 | -1.071 | -0.305 |

Based on estimated marginal means

*. The mean difference is significant at the 0.05 level.

b. Adjustment for multiple comparisons: Least Significant Difference (equivalent to no adjustments).

HC=0, MA use=1, MA dependence=2, MAP=3

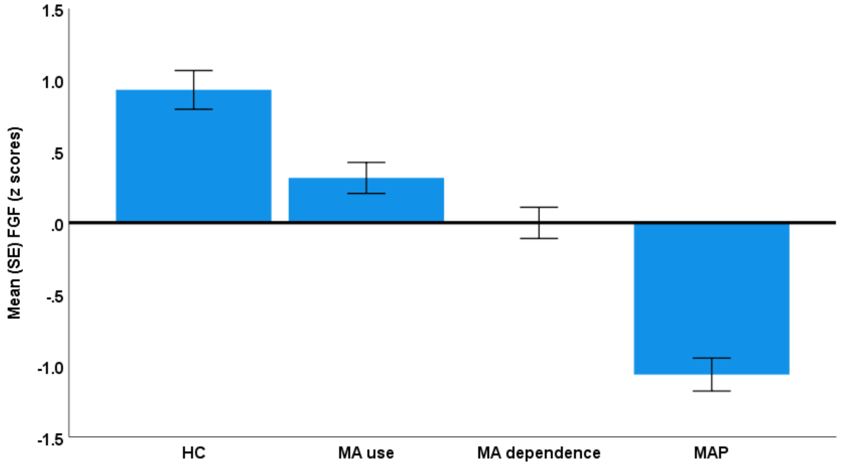


Figure S3. FGF levels in healthy controls (HC) and people with methamphetamine (MA) use, MA dependence, and MA-induced psychosis (MAP) ($F=47.71$, $df=3/164$, $p<0.001$).

**Pairwise Comparisons**

Dependent Variable: FGF

| (I) HC_Abuse_Dependence_Psychosis | (J) HC_Abuse_Dependence_Psychosis | Mean Difference (I-J) | Std. Error | Sig.[b] | 95% Confidence Interval for Difference[b] | |
|---|---|---|---|---|---|---|
| | | | | | Lower Bound | Upper Bound |
| 0 | 1 | .617* | 0.178 | 0.001 | 0.264 | 0.969 |
| | 2 | .932* | 0.177 | 0.000 | 0.583 | 1.280 |
| | 3 | 1.994* | 0.176 | 0.000 | 1.647 | 2.341 |
| 1 | 0 | -.617* | 0.178 | 0.001 | -0.969 | -0.264 |
| | 2 | .315* | 0.154 | 0.042 | 0.011 | 0.619 |
| | 3 | 1.377* | 0.163 | 0.000 | 1.055 | 1.699 |
| 2 | 0 | -.932* | 0.177 | 0.000 | -1.280 | -0.583 |
| | 1 | -.315* | 0.154 | 0.042 | -0.619 | -0.011 |
| | 3 | 1.062* | 0.160 | 0.000 | 0.747 | 1.378 |
| 3 | 0 | -1.994* | 0.176 | 0.000 | -2.341 | -1.647 |
| | 1 | -1.377* | 0.163 | 0.000 | -1.699 | -1.055 |
| | 2 | -1.062* | 0.160 | 0.000 | -1.378 | -0.747 |

Based on estimated marginal means

*. The mean difference is significant at the 0.05 level.

b. Adjustment for multiple comparisons: Least Significant Difference (equivalent to no adjustments).

HC=0, MA use=1, MA dependence=2, MAP=3

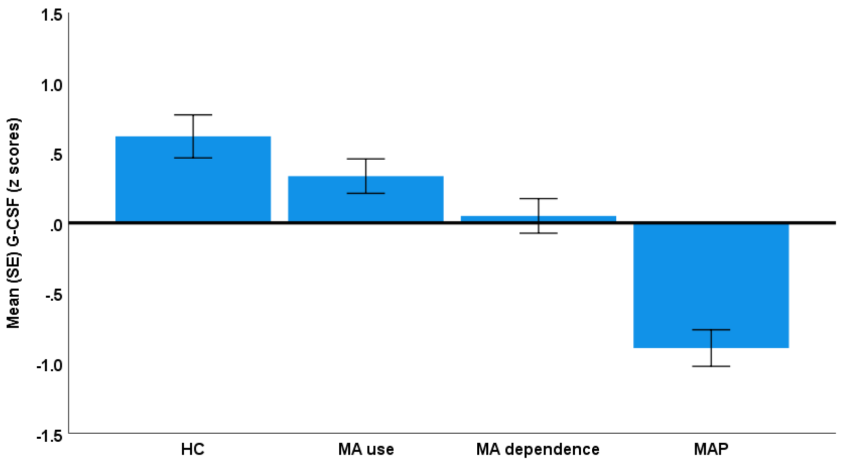


Figure S4. G-CSF levels in healthy controls (HC) and people with methamphetamine (MA) use, MA dependence, and MA-induced psychosis (MAP) (F=23.77, df=3/164, p<0.001).

**Pairwise Comparisons**

Dependent Variable: G-CSF

| (I) HC_Abuse_ Dependence_ Psychosis | (J) HC_Abuse_ Dependence_ Psychosis | Mean Difference (I-J) | Std. Error | Sig.[b] | 95% Confidence Interval for Difference[b] | |
|---|---|---|---|---|---|---|
| | | | | | Lower Bound | Upper Bound |
| 0 | 1 | 0.283 | 0.202 | 0.162 | -0.115 | 0.681 |
| | 2 | .567* | 0.200 | 0.005 | 0.173 | 0.961 |
| | 3 | 1.509* | 0.198 | 0.000 | 1.118 | 1.901 |
| 1 | 0 | -0.283 | 0.202 | 0.162 | -0.681 | 0.115 |
| | 2 | 0.284 | 0.174 | 0.105 | -0.060 | 0.627 |
| | 3 | 1.226* | 0.184 | 0.000 | 0.862 | 1.590 |
| 2 | 0 | -.567* | 0.200 | 0.005 | -0.961 | -0.173 |
| | 1 | -0.284 | 0.174 | 0.105 | -0.627 | 0.060 |
| | 3 | .942* | 0.181 | 0.000 | 0.586 | 1.299 |
| 3 | 0 | -1.509* | 0.198 | 0.000 | -1.901 | -1.118 |
| | 1 | -1.226* | 0.184 | 0.000 | -1.590 | -0.862 |
| | 2 | -.942* | 0.181 | 0.000 | -1.299 | -0.586 |

Based on estimated marginal means

*. The mean difference is significant at the 0.05 level.

b. Adjustment for multiple comparisons: Least Significant Difference (equivalent to no adjustments).

HC=0, MA use=1, MA dependence=2, MAP=3

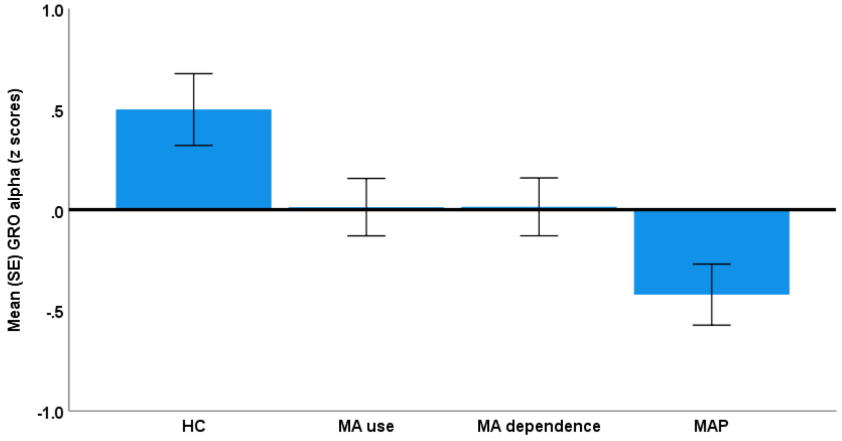


Figure S5. CXCL1 (GRO-α) levels in healthy controls (HC) and people with methamphetamine (MA) use, MA dependence, and MA-induced psychosis (MAP) (F=5.33, df=3/164, p=0.002).

**Pairwise Comparisons**

Dependent Variable: GRO-α

| (I) HC_Abuse_ Dependence_ Psychosis | (J) HC_Abuse_ Dependence_ Psychosis | Mean Difference (I-J) | Std. Error | Sig.[b] | 95% Confidence Interval for Difference[b] | |
|---|---|---|---|---|---|---|
| | | | | | Lower Bound | Upper Bound |
| 0 | 1 | .485* | 0.235 | 0.040 | 0.022 | 0.949 |
| | 2 | .484* | 0.233 | 0.039 | 0.025 | 0.943 |
| | 3 | .920* | 0.231 | 0.000 | 0.464 | 1.377 |
| 1 | 0 | -.485* | 0.235 | 0.040 | -0.949 | -0.022 |
| | 2 | -0.002 | 0.202 | 0.994 | -0.401 | 0.398 |
| | 3 | .435* | 0.215 | 0.044 | 0.011 | 0.859 |
| 2 | 0 | -.484* | 0.233 | 0.039 | -0.943 | -0.025 |
| | 1 | 0.002 | 0.202 | 0.994 | -0.398 | 0.401 |
| | 3 | .437* | 0.210 | 0.040 | 0.021 | 0.852 |
| 3 | 0 | -.920* | 0.231 | 0.000 | -1.377 | -0.464 |
| | 1 | -.435* | 0.215 | 0.044 | -0.859 | -0.011 |
| | 2 | -.437* | 0.210 | 0.040 | -0.852 | -0.021 |

Based on estimated marginal means

*. The mean difference is significant at the 0.05 level.

b. Adjustment for multiple comparisons: Least Significant Difference (equivalent to no adjustments).

HC=0, MA use=1, MA dependence=2, MAP=3

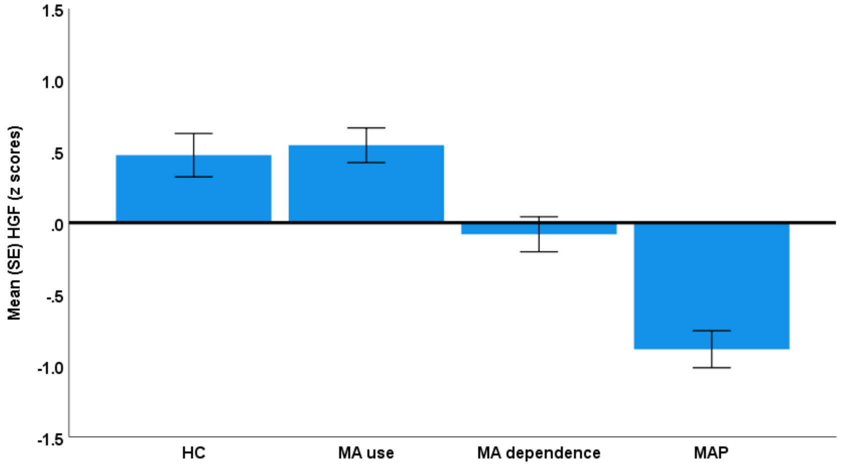


Figure S6. HGF levels in healthy controls (HC) and people with methamphetamine (MA) use, MA dependence, and MA-induced psychosis (MAP) (F=25.43, df=3/164, p<0.001).

**Pairwise Comparisons**

Dependent Variable: HGF

| (I) HC_Abuse_ Dependence_ Psychosis | (J) HC_Abuse_ Dependence_ Psychosis | Mean Difference (I-J) | Std. Error | Sig.[b] | 95% Confidence Interval for Difference[b] | |
|---|---|---|---|---|---|---|
| | | | | | Lower Bound | Upper Bound |
| 0 | 1 | -0.070 | 0.200 | 0.728 | -0.464 | 0.325 |
| | 2 | .554* | 0.198 | 0.006 | 0.163 | 0.945 |
| | 3 | 1.359* | 0.197 | 0.000 | 0.971 | 1.748 |
| 1 | 0 | 0.070 | 0.200 | 0.728 | -0.325 | 0.464 |
| | 2 | .624* | 0.172 | 0.000 | 0.283 | 0.964 |
| | 3 | 1.429* | 0.183 | 0.000 | 1.068 | 1.790 |
| 2 | 0 | -.554* | 0.198 | 0.006 | -0.945 | -0.163 |
| | 1 | -.624* | 0.172 | 0.000 | -0.964 | -0.283 |
| | 3 | .805* | 0.179 | 0.000 | 0.452 | 1.159 |
| 3 | 0 | -1.359* | 0.197 | 0.000 | -1.748 | -0.971 |
| | 1 | -1.429* | 0.183 | 0.000 | -1.790 | -1.068 |
| | 2 | -.805* | 0.179 | 0.000 | -1.159 | -0.452 |

Based on estimated marginal means

*. The mean difference is significant at the 0.05 level.

b. Adjustment for multiple comparisons: Least Significant Difference (equivalent to no adjustments).

HC=0, MA use=1, MA dependence=2, MAP=3

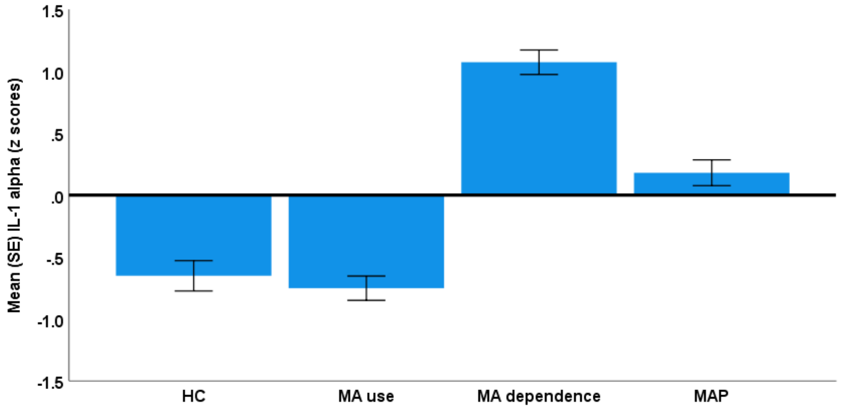


Figure S7. IL-1α levels in healthy controls (HC) and people with methamphetamine (MA) use, MA dependence, and MA-induced psychosis (MAP) ($F$=69.76, df=3/164, $p$<0.001).

**Pairwise Comparisons**

Dependent Variable: IL-1α

| (I) HC_Abuse_ Dependence_ Psychosis | (J) HC_Abuse_ Dependence_ Psychosis | Mean Difference (I-J) | Std. Error | Sig.[b] | 95% Confidence Interval for Difference[b] | |
|---|---|---|---|---|---|---|
| | | | | | Lower Bound | Upper Bound |
| 0 | 1 | 0.100 | 0.161 | 0.534 | -0.218 | 0.419 |
| | 2 | -1.725* | 0.160 | 0.000 | -2.040 | -1.410 |
| | 3 | -.832* | 0.159 | 0.000 | -1.145 | -0.519 |
| 1 | 0 | -0.100 | 0.161 | 0.534 | -0.419 | 0.218 |
| | 2 | -1.825* | 0.139 | 0.000 | -2.100 | -1.551 |
| | 3 | -.933* | 0.147 | 0.000 | -1.224 | -0.642 |
| 2 | 0 | 1.725* | 0.160 | 0.000 | 1.410 | 2.040 |
| | 1 | 1.825* | 0.139 | 0.000 | 1.551 | 2.100 |
| | 3 | .892* | 0.144 | 0.000 | 0.607 | 1.178 |
| 3 | 0 | .832* | 0.159 | 0.000 | 0.519 | 1.145 |
| | 1 | .933* | 0.147 | 0.000 | 0.642 | 1.224 |
| | 2 | -.892* | 0.144 | 0.000 | -1.178 | -0.607 |

Based on estimated marginal means

*. The mean difference is significant at the 0.05 level.

b. Adjustment for multiple comparisons: Least Significant Difference (equivalent to no adjustments).

HC=0, MA use=1, MA dependence=2, MAP=3

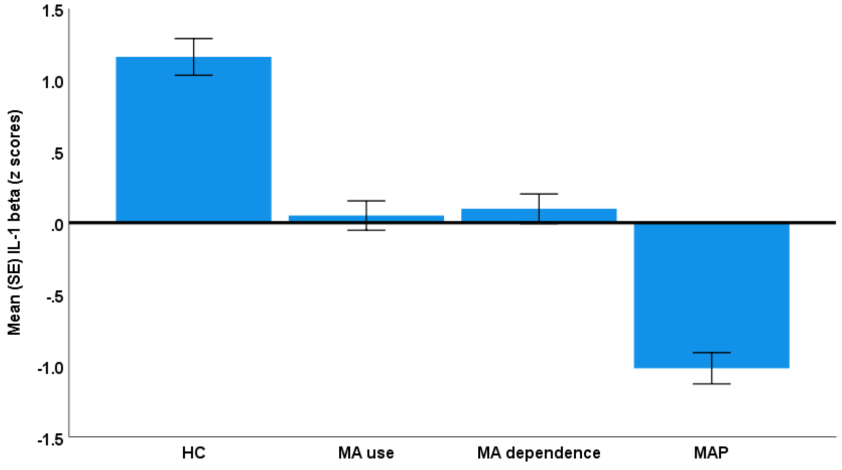


Figure S8. IL-1β levels in healthy controls (HC) and people with methamphetamine (MA) use, MA dependence, and MA-induced psychosis (MAP) ($F=58.11$, $df=3/164$, $p<0.001$).

**Pairwise Comparisons**

Dependent Variable: IL-1β

| (I) HC_Abuse_Dependence_Psychosis | (J) HC_Abuse_Dependence_Psychosis | Mean Difference (I-J) | Std. Error | Sig.[b] | 95% Confidence Interval for Difference[b] Lower Bound | Upper Bound |
|---|---|---|---|---|---|---|
| 0 | 1 | 1.111* | 0.169 | 0.000 | 0.777 | 1.446 |
| | 2 | 1.064* | 0.168 | 0.000 | 0.733 | 1.395 |
| | 3 | 2.181* | 0.167 | 0.000 | 1.852 | 2.510 |
| 1 | 0 | -1.111* | 0.169 | 0.000 | -1.446 | -0.777 |
| | 2 | -0.047 | 0.146 | 0.746 | -0.336 | 0.241 |
| | 3 | 1.070* | 0.155 | 0.000 | 0.764 | 1.375 |
| 2 | 0 | -1.064* | 0.168 | 0.000 | -1.395 | -0.733 |
| | 1 | 0.047 | 0.146 | 0.746 | -0.241 | 0.336 |
| | 3 | 1.117* | 0.152 | 0.000 | 0.817 | 1.416 |
| 3 | 0 | -2.181* | 0.167 | 0.000 | -2.510 | -1.852 |
| | 1 | -1.070* | 0.155 | 0.000 | -1.375 | -0.764 |
| | 2 | -1.117* | 0.152 | 0.000 | -1.416 | -0.817 |

Based on estimated marginal means

*. The mean difference is significant at the 0.05 level.

b. Adjustment for multiple comparisons: Least Significant Difference (equivalent to no adjustments).

HC=0, MA use=1, MA dependence=2, MAP=3

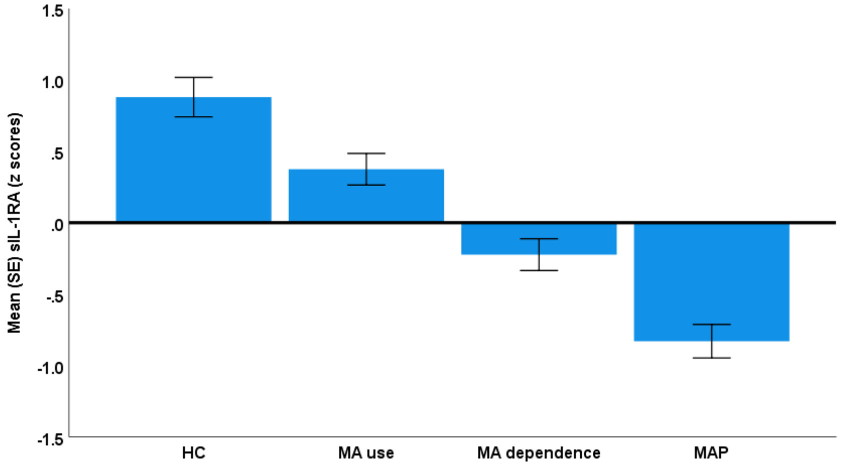


Figure S9. sIL-1RA levels in healthy controls (HC) and people with methamphetamine (MA) use, MA dependence, and MA-induced psychosis (MAP) (F=35.87, df=3/164, p<0.001).

**Pairwise Comparisons**

Dependent Variable: sIL-1RA

| (I) HC_Abuse_ Dependence_ Psychosis | (J) HC_Abuse_ Dependence_ Psychosis | Mean Difference (I-J) | Std. Error | Sig.[b] | 95% Confidence Interval for Difference[b] | |
|---|---|---|---|---|---|---|
| | | | | | Lower Bound | Upper Bound |
| 0 | 1 | .505* | 0.182 | 0.006 | 0.146 | 0.864 |
| | 2 | 1.103* | 0.180 | 0.000 | 0.748 | 1.459 |
| | 3 | 1.709* | 0.179 | 0.000 | 1.356 | 2.062 |
| 1 | 0 | -.505* | 0.182 | 0.006 | -0.864 | -0.146 |
| | 2 | .598* | 0.157 | 0.000 | 0.289 | 0.908 |
| | 3 | 1.204* | 0.166 | 0.000 | 0.876 | 1.533 |
| 2 | 0 | -1.103* | 0.180 | 0.000 | -1.459 | -0.748 |
| | 1 | -.598* | 0.157 | 0.000 | -0.908 | -0.289 |
| | 3 | .606* | 0.163 | 0.000 | 0.284 | 0.927 |
| 3 | 0 | -1.709* | 0.179 | 0.000 | -2.062 | -1.356 |
| | 1 | -1.204* | 0.166 | 0.000 | -1.533 | -0.876 |
| | 2 | -.606* | 0.163 | 0.000 | -0.927 | -0.284 |

Based on estimated marginal means

*. The mean difference is significant at the 0.05 level.

b. Adjustment for multiple comparisons: Least Significant Difference (equivalent to no adjustments).

HC=0, MA use=1, MA dependence=2, MAP=3

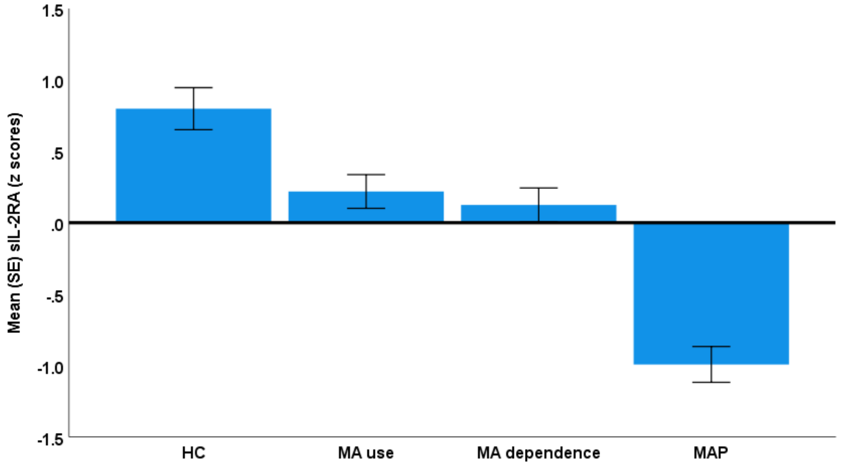


Figure S10. sIL-2RA levels in healthy controls (HC) and people with methamphetamine (MA) use, MA dependence, and MA-induced psychosis (MAP) (F=32.87, df=3/164, p<0.001).

**Pairwise Comparisons**

Dependent Variable: sIL-2RA

| (I) HC_Abuse_ Dependence_ Psychosis | (J) HC_Abuse_ Dependence_ Psychosis | Mean Difference (I-J) | Std. Error | Sig.[b] | 95% Confidence Interval for Difference[b] | |
|---|---|---|---|---|---|---|
| | | | | | Lower Bound | Upper Bound |
| 0 | 1 | .580* | 0.194 | 0.003 | 0.197 | 0.963 |
| | 2 | .674* | 0.192 | 0.001 | 0.295 | 1.053 |
| | 3 | 1.792* | 0.191 | 0.000 | 1.415 | 2.168 |
| 1 | 0 | -.580* | 0.194 | 0.003 | -0.963 | -0.197 |
| | 2 | 0.094 | 0.167 | 0.575 | -0.236 | 0.424 |
| | 3 | 1.211* | 0.177 | 0.000 | 0.861 | 1.562 |
| 2 | 0 | -.674* | 0.192 | 0.001 | -1.053 | -0.295 |
| | 1 | -0.094 | 0.167 | 0.575 | -0.424 | 0.236 |
| | 3 | 1.117* | 0.174 | 0.000 | 0.774 | 1.460 |
| 3 | 0 | -1.792* | 0.191 | 0.000 | -2.168 | -1.415 |
| | 1 | -1.211* | 0.177 | 0.000 | -1.562 | -0.861 |
| | 2 | -1.117* | 0.174 | 0.000 | -1.460 | -0.774 |

Based on estimated marginal means

*. The mean difference is significant at the 0.05 level.

b. Adjustment for multiple comparisons: Least Significant Difference (equivalent to no adjustments).

HC=0, MA use=1, MA dependence=2, MAP=3

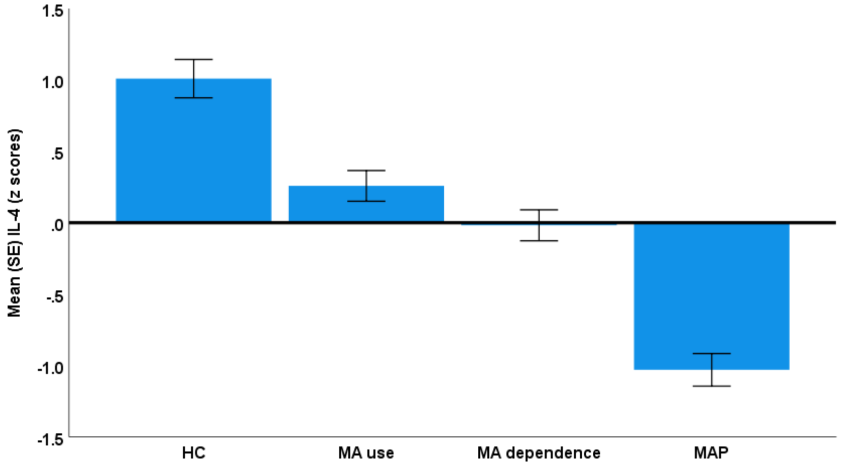


Figure S11. IL-4 levels in healthy controls (HC) and people with methamphetamine (MA) use, MA dependence, and MA-induced psychosis (MAP) ($F=48.96$, $df=3/164$, $p<0.001$).

**Pairwise Comparisons**

Dependent Variable: IL-4

| (I) HC_Abuse_Dependence_Psychosis | (J) HC_Abuse_Dependence_Psychosis | Mean Difference (I-J) | Std. Error | Sig.[b] | 95% Confidence Interval for Difference[b] | |
|---|---|---|---|---|---|---|
| | | | | | Lower Bound | Upper Bound |
| 0 | 1 | .751* | 0.177 | 0.000 | 0.402 | 1.100 |
| | 2 | 1.027* | 0.175 | 0.000 | 0.682 | 1.373 |
| | 3 | 2.040* | 0.174 | 0.000 | 1.696 | 2.383 |
| 1 | 0 | -.751* | 0.177 | 0.000 | -1.100 | -0.402 |
| | 2 | 0.276 | 0.152 | 0.072 | -0.025 | 0.577 |
| | 3 | 1.289* | 0.162 | 0.000 | 0.969 | 1.608 |
| 2 | 0 | -1.027* | 0.175 | 0.000 | -1.373 | -0.682 |
| | 1 | -0.276 | 0.152 | 0.072 | -0.577 | 0.025 |
| | 3 | 1.013* | 0.158 | 0.000 | 0.700 | 1.325 |
| 3 | 0 | -2.040* | 0.174 | 0.000 | -2.383 | -1.696 |
| | 1 | -1.289* | 0.162 | 0.000 | -1.608 | -0.969 |
| | 2 | -1.013* | 0.158 | 0.000 | -1.325 | -0.700 |

Based on estimated marginal means

*. The mean difference is significant at the 0.05 level.

b. Adjustment for multiple comparisons: Least Significant Difference (equivalent to no adjustments).

HC=0, MA use=1, MA dependence=2, MAP=3

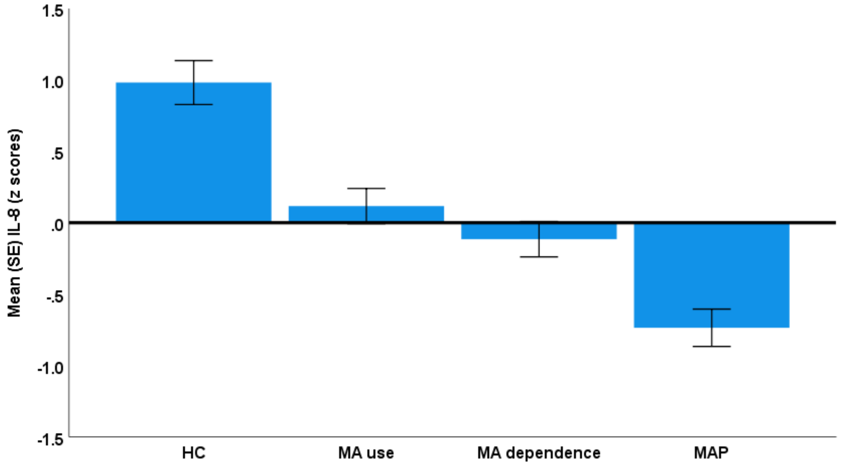


Figure S12.CXCL8 or IL-8 levels in healthy controls (HC) and people with methamphetamine (MA) use, MA dependence, and MA-induced psychosis (MAP) (F=25.29, df=3/164, p<0.001).

**Pairwise Comparisons**

Dependent Variable: IL-8

| (I) HC_Abuse_Dependence_Psychosis | (J) HC_Abuse_Dependence_Psychosis | Mean Difference (I-J) | Std. Error | Sig.[b] | 95% Confidence Interval for Difference[b] | |
|---|---|---|---|---|---|---|
| | | | | | Lower Bound | Upper Bound |
| 0 | 1 | .865* | 0.203 | 0.000 | 0.465 | 1.265 |
| | 2 | 1.097* | 0.201 | 0.000 | 0.701 | 1.493 |
| | 3 | 1.717* | 0.199 | 0.000 | 1.324 | 2.111 |
| 1 | 0 | -.865* | 0.203 | 0.000 | -1.265 | -0.465 |
| | 2 | 0.232 | 0.175 | 0.186 | -0.113 | 0.577 |
| | 3 | .852* | 0.185 | 0.000 | 0.487 | 1.218 |
| 2 | 0 | -1.097* | 0.201 | 0.000 | -1.493 | -0.701 |
| | 1 | -0.232 | 0.175 | 0.186 | -0.577 | 0.113 |
| | 3 | .621* | 0.181 | 0.001 | 0.262 | 0.979 |
| 3 | 0 | -1.717* | 0.199 | 0.000 | -2.111 | -1.324 |
| | 1 | -.852* | 0.185 | 0.000 | -1.218 | -0.487 |
| | 2 | -.621* | 0.181 | 0.001 | -0.979 | -0.262 |

Based on estimated marginal means

*. The mean difference is significant at the 0.05 level.

b. Adjustment for multiple comparisons: Least Significant Difference (equivalent to no adjustments).

HC=0, MA use=1, MA dependence=2, MAP=3

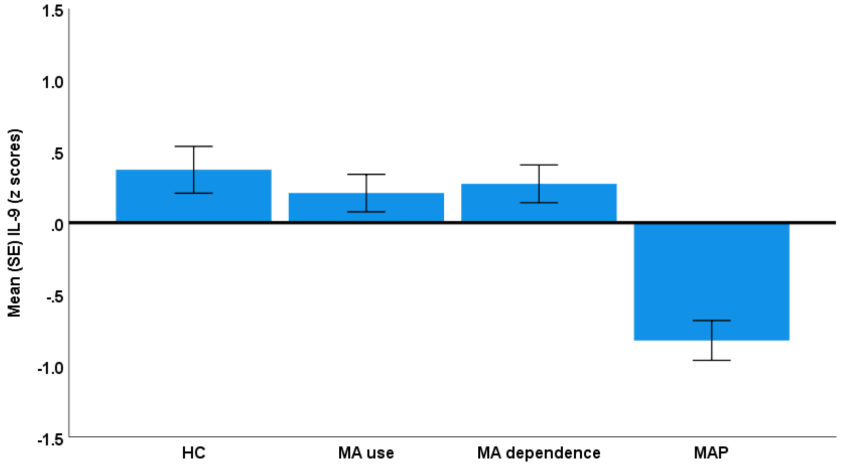


Figure S13. IL-9 levels in healthy controls (HC) and people with methamphetamine (MA) use, MA dependence, and MA-induced psychosis (MAP) (F=15.67, df=3/164, p<0.001).

**Pairwise Comparisons**

Dependent Variable: IL-9

| (I) HC_Abuse_ Dependence_ Psychosis | (J) HC_Abuse_ Dependence_ Psychosis | Mean Difference (I-J) | Std. Error | Sig.[b] | 95% Confidence Interval for Difference[b] | |
|---|---|---|---|---|---|---|
| | | | | | Lower Bound | Upper Bound |
| 0 | 1 | 0.163 | 0.216 | 0.451 | -0.263 | 0.589 |
| | 2 | 0.098 | 0.214 | 0.646 | -0.323 | 0.520 |
| | 3 | 1.195* | 0.212 | 0.000 | 0.776 | 1.614 |
| 1 | 0 | -0.163 | 0.216 | 0.451 | -0.589 | 0.263 |
| | 2 | -0.065 | 0.186 | 0.728 | -0.432 | 0.302 |
| | 3 | 1.032* | 0.197 | 0.000 | 0.643 | 1.422 |
| 2 | 0 | -0.098 | 0.214 | 0.646 | -0.520 | 0.323 |
| | 1 | 0.065 | 0.186 | 0.728 | -0.302 | 0.432 |
| | 3 | 1.097* | 0.193 | 0.000 | 0.716 | 1.478 |
| 3 | 0 | -1.195* | 0.212 | 0.000 | -1.614 | -0.776 |
| | 1 | -1.032* | 0.197 | 0.000 | -1.422 | -0.643 |
| | 2 | -1.097* | 0.193 | 0.000 | -1.478 | -0.716 |

Based on estimated marginal means

*. The mean difference is significant at the 0.05 level.

b. Adjustment for multiple comparisons: Least Significant Difference (equivalent to no adjustments).

HC=0, MA use=1, MA dependence=2, MAP=3

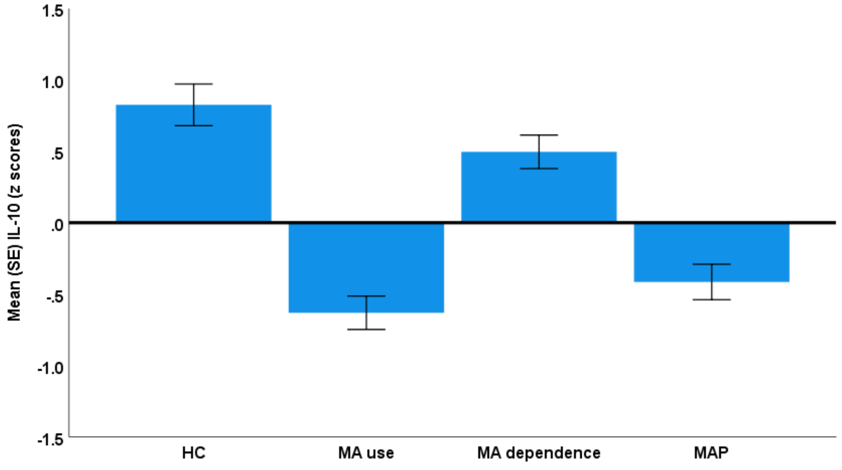


Figure S14. IL-10 levels in healthy controls (HC) and people with methamphetamine (MA) use, MA dependence, and MA-induced psychosis (MAP) (F=30.46, df=3/164, p<0.001).

**Pairwise Comparisons**

Dependent Variable: IL-10

| (I) HC_Abuse_ Dependence_ Psychosis | (J) HC_Abuse_ Dependence_ Psychosis | Mean Difference (I-J) | Std. Error | Sig.[b] | 95% Confidence Interval for Difference[b] | |
|---|---|---|---|---|---|---|
| | | | | | Lower Bound | Upper Bound |
| 0 | 1 | 1.456* | 0.192 | 0.000 | 1.077 | 1.835 |
| | 2 | 0.329 | 0.190 | 0.086 | -0.047 | 0.704 |
| | 3 | 1.240* | 0.189 | 0.000 | 0.867 | 1.614 |
| 1 | 0 | -1.456* | 0.192 | 0.000 | -1.835 | -1.077 |
| | 2 | -1.127* | 0.166 | 0.000 | -1.454 | -0.800 |
| | 3 | -0.216 | 0.176 | 0.221 | -0.562 | 0.131 |
| 2 | 0 | -0.329 | 0.190 | 0.086 | -0.704 | 0.047 |
| | 1 | 1.127* | 0.166 | 0.000 | 0.800 | 1.454 |
| | 3 | .912* | 0.172 | 0.000 | 0.572 | 1.251 |
| 3 | 0 | -1.240* | 0.189 | 0.000 | -1.614 | -0.867 |
| | 1 | 0.216 | 0.176 | 0.221 | -0.131 | 0.562 |
| | 2 | -.912* | 0.172 | 0.000 | -1.251 | -0.572 |

Based on estimated marginal means

*. The mean difference is significant at the 0.05 level.

b. Adjustment for multiple comparisons: Least Significant Difference (equivalent to no adjustments).

HC=0, MA use=1, MA dependence=2, MAP=3

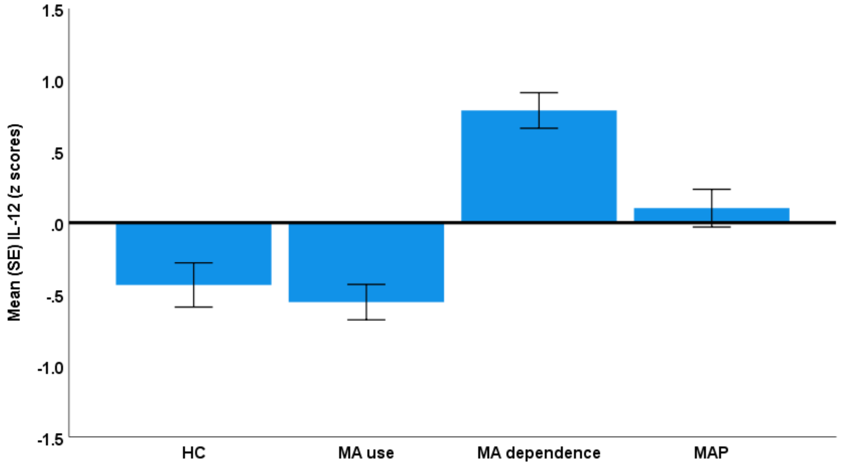


Figure S15. IL-12 p70 levels in healthy controls (HC) and people with methamphetamine (MA) use, MA dependence, and MA-induced psychosis (MAP) (F=22.87, df=3/164, p<0.001).

**Pairwise Comparisons**

Dependent Variable: IL-12p70

| (I) HC_Abuse_ Dependence_ Psychosis | (J) HC_Abuse_ Dependence_ Psychosis | Mean Difference (I-J) | Std. Error | Sig.[b] | 95% Confidence Interval for Difference[b] | |
|---|---|---|---|---|---|---|
| | | | | | Lower Bound | Upper Bound |
| 0 | 1 | 0.120 | 0.204 | 0.556 | -0.282 | 0.523 |
| | 2 | -1.222* | 0.202 | 0.000 | -1.621 | -0.823 |
| | 3 | -.538* | 0.201 | 0.008 | -0.934 | -0.142 |
| 1 | 0 | -0.120 | 0.204 | 0.556 | -0.523 | 0.282 |
| | 2 | -1.343* | 0.176 | 0.000 | -1.690 | -0.995 |
| | 3 | -.658* | 0.186 | 0.001 | -1.026 | -0.290 |
| 2 | 0 | 1.222* | 0.202 | 0.000 | 0.823 | 1.621 |
| | 1 | 1.343* | 0.176 | 0.000 | 0.995 | 1.690 |
| | 3 | .684* | 0.183 | 0.000 | 0.324 | 1.045 |
| 3 | 0 | .538* | 0.201 | 0.008 | 0.142 | 0.934 |
| | 1 | .658* | 0.186 | 0.001 | 0.290 | 1.026 |
| | 2 | -.684* | 0.183 | 0.000 | -1.045 | -0.324 |

Based on estimated marginal means

*. The mean difference is significant at the 0.05 level.

b. Adjustment for multiple comparisons: Least Significant Difference (equivalent to no adjustments).

HC=0, MA use=1, MA dependence=2, MAP=3

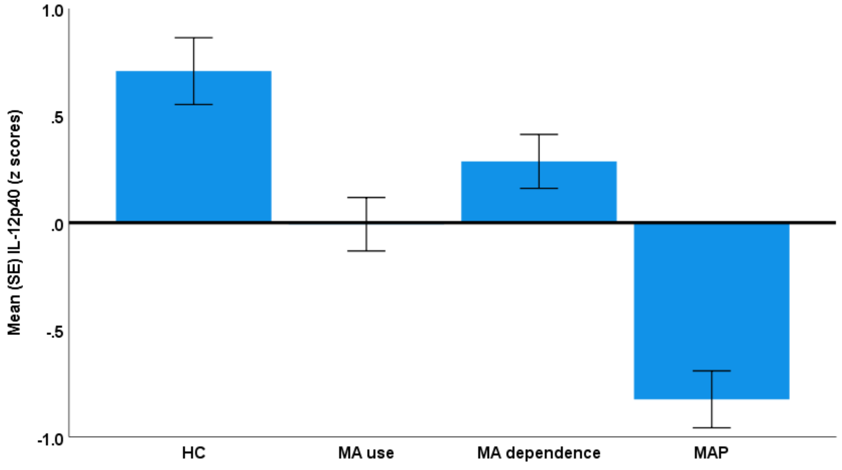


Figure S16. IL-12p40 levels in healthy controls (HC) and people with methamphetamine (MA) use, MA dependence, and MA-induced psychosis (MAP) (F=22.09, df=3/164, p<0.001).

**Pairwise Comparisons**

Dependent Variable: IL-12p40

| (I) HC_Abuse_ Dependence_ Psychosis | (J) HC_Abuse_ Dependence_ Psychosis | Mean Difference (I-J) | Std. Error | Sig.[b] | 95% Confidence Interval for Difference[b] | |
|---|---|---|---|---|---|---|
| | | | | | Lower Bound | Upper Bound |
| 0 | 1 | .715* | 0.205 | 0.001 | 0.309 | 1.120 |
| | 2 | .421* | 0.203 | 0.040 | 0.020 | 0.823 |
| | 3 | 1.533* | 0.202 | 0.000 | 1.133 | 1.932 |
| 1 | 0 | -.715* | 0.205 | 0.001 | -1.120 | -0.309 |
| | 2 | -0.293 | 0.177 | 0.100 | -0.643 | 0.056 |
| | 3 | .818* | 0.188 | 0.000 | 0.447 | 1.189 |
| 2 | 0 | -.421* | 0.203 | 0.040 | -0.823 | -0.020 |
| | 1 | 0.293 | 0.177 | 0.100 | -0.056 | 0.643 |
| | 3 | 1.111* | 0.184 | 0.000 | 0.748 | 1.475 |
| 3 | 0 | -1.533* | 0.202 | 0.000 | -1.932 | -1.133 |
| | 1 | -.818* | 0.188 | 0.000 | -1.189 | -0.447 |
| | 2 | -1.111* | 0.184 | 0.000 | -1.475 | -0.748 |

Based on estimated marginal means

*. The mean difference is significant at the 0.05 level.

b. Adjustment for multiple comparisons: Least Significant Difference (equivalent to no adjustments).

HC=0, MA use=1, MA dependence=2, MAP=3

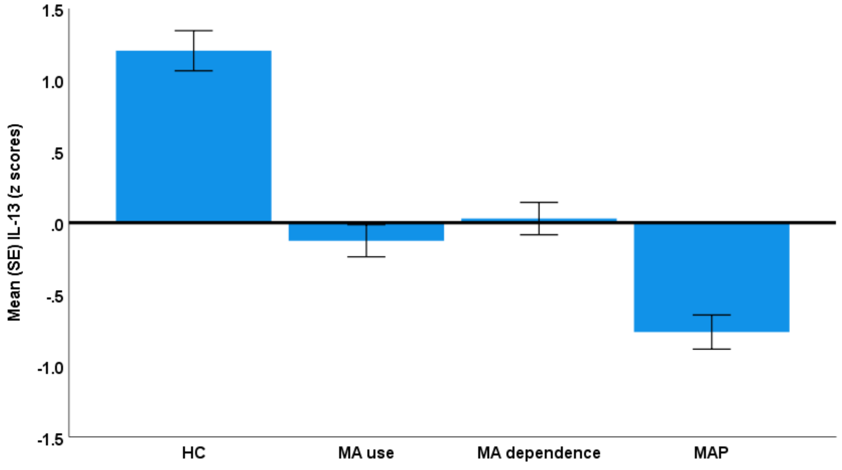


Figure S17. IL-13 levels in healthy controls (HC) and people with methamphetamine (MA) use, MA dependence, and MA-induced psychosis (MAP) ($F=39.64$, $df=3/164$, $p<0.001$).

**Pairwise Comparisons**

Dependent Variable: IL-13

| (I) HC_Abuse_ Dependence_ Psychosis | (J) HC_Abuse_ Dependence_ Psychosis | Mean Difference (I-J) | Std. Error | Sig.[b] | 95% Confidence Interval for Difference[b] | |
|---|---|---|---|---|---|---|
| | | | | | Lower Bound | Upper Bound |
| 0 | 1 | 1.331* | 0.185 | 0.000 | 0.966 | 1.696 |
| | 2 | 1.175* | 0.183 | 0.000 | 0.814 | 1.537 |
| | 3 | 1.970* | 0.182 | 0.000 | 1.611 | 2.330 |
| 1 | 0 | -1.331* | 0.185 | 0.000 | -1.696 | -0.966 |
| | 2 | -0.156 | 0.159 | 0.331 | -0.470 | 0.159 |
| | 3 | .640* | 0.169 | 0.000 | 0.306 | 0.973 |
| 2 | 0 | -1.175* | 0.183 | 0.000 | -1.537 | -0.814 |
| | 1 | 0.156 | 0.159 | 0.331 | -0.159 | 0.470 |
| | 3 | .795* | 0.166 | 0.000 | 0.468 | 1.122 |
| 3 | 0 | -1.970* | 0.182 | 0.000 | -2.330 | -1.611 |
| | 1 | -.640* | 0.169 | 0.000 | -0.973 | -0.306 |
| | 2 | -.795* | 0.166 | 0.000 | -1.122 | -0.468 |

Based on estimated marginal means

*. The mean difference is significant at the 0.05 level.

b. Adjustment for multiple comparisons: Least Significant Difference (equivalent to no adjustments).

HC=0, MA use=1, MA dependence=2, MAP=3

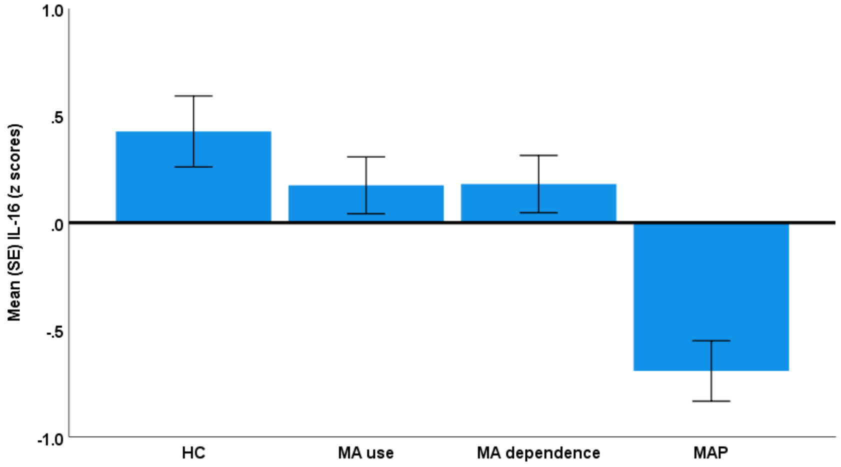


Figure S18. IL-16 levels in healthy controls (HC) and people with methamphetamine (MA) use, MA dependence, and MA-induced psychosis (MAP) (F=11.56, df=3/164, p<0.001).

**Pairwise Comparisons**

Dependent Variable: IL-16

| (I) HC_Abuse_ Dependence_ Psychosis | (J) HC_Abuse_ Dependence_ Psychosis | Mean Difference (I-J) | Std. Error | Sig.[b] | 95% Confidence Interval for Difference[b] | |
|---|---|---|---|---|---|---|
| | | | | | Lower Bound | Upper Bound |
| 0 | 1 | 0.251 | 0.218 | 0.251 | -0.179 | 0.682 |
| | 2 | 0.245 | 0.216 | 0.257 | -0.181 | 0.672 |
| | 3 | 1.118* | 0.215 | 0.000 | 0.694 | 1.542 |
| 1 | 0 | -0.251 | 0.218 | 0.251 | -0.682 | 0.179 |
| | 2 | -0.006 | 0.188 | 0.975 | -0.377 | 0.365 |
| | 3 | .867* | 0.199 | 0.000 | 0.473 | 1.260 |
| 2 | 0 | -0.245 | 0.216 | 0.257 | -0.672 | 0.181 |
| | 1 | 0.006 | 0.188 | 0.975 | -0.365 | 0.377 |
| | 3 | .873* | 0.195 | 0.000 | 0.487 | 1.258 |
| 3 | 0 | -1.118* | 0.215 | 0.000 | -1.542 | -0.694 |
| | 1 | -.867* | 0.199 | 0.000 | -1.260 | -0.473 |
| | 2 | -.873* | 0.195 | 0.000 | -1.258 | -0.487 |

Based on estimated marginal means

*. The mean difference is significant at the 0.05 level.

b. Adjustment for multiple comparisons: Least Significant Difference (equivalent to no adjustments).

HC=0, MA use=1, MA dependence=2, MAP=3

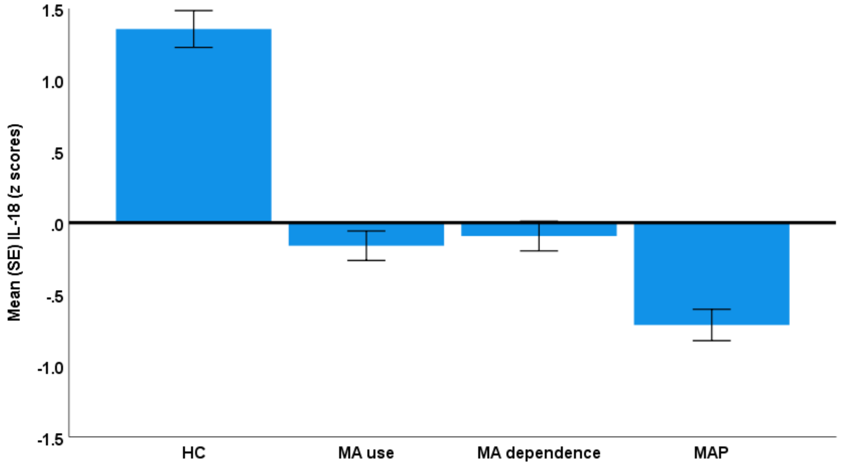


Figure S19. IL-18 levels in healthy controls (HC) and people with methamphetamine (MA) use, MA dependence, and MA-induced psychosis (MAP) (F=53.10, df=3/164, p<0.001).

**Pairwise Comparisons**

Dependent Variable: IL-18

| (I) HC_Abuse_ Dependence_ Psychosis | (J) HC_Abuse_ Dependence_ Psychosis | Mean Difference (I-J) | Std. Error | Sig.[b] | 95% Confidence Interval for Difference[b] | |
|---|---|---|---|---|---|---|
| | | | | | Lower Bound | Upper Bound |
| 0 | 1 | 1.518* | 0.170 | 0.000 | 1.183 | 1.853 |
| | 2 | 1.450* | 0.168 | 0.000 | 1.118 | 1.782 |
| | 3 | 2.073* | 0.167 | 0.000 | 1.743 | 2.403 |
| 1 | 0 | -1.518* | 0.170 | 0.000 | -1.853 | -1.183 |
| | 2 | -0.068 | 0.146 | 0.643 | -0.357 | 0.221 |
| | 3 | .555* | 0.155 | 0.000 | 0.248 | 0.862 |
| 2 | 0 | -1.450* | 0.168 | 0.000 | -1.782 | -1.118 |
| | 1 | 0.068 | 0.146 | 0.643 | -0.221 | 0.357 |
| | 3 | .623* | 0.152 | 0.000 | 0.323 | 0.923 |
| 3 | 0 | -2.073* | 0.167 | 0.000 | -2.403 | -1.743 |
| | 1 | -.555* | 0.155 | 0.000 | -0.862 | -0.248 |
| | 2 | -.623* | 0.152 | 0.000 | -0.923 | -0.323 |

Based on estimated marginal means

*. The mean difference is significant at the 0.05 level.

b. Adjustment for multiple comparisons: Least Significant Difference (equivalent to no adjustments).

HC=0, MA use=1, MA dependence=2, MAP=3

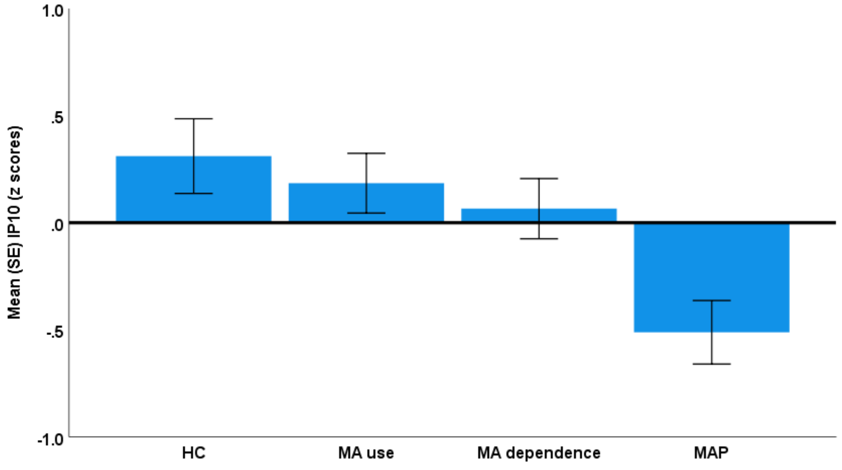


Figure S20. CXCL10 or IP10 levels in healthy controls (HC) and people with methamphetamine (MA) use, MA dependence, and MA-induced psychosis (MAP) (F=5.72, df=3/164, p<0.001).

**Pairwise Comparisons**

Dependent Variable: IP10

| (I) HC_Abuse_Dependence_Psychosis | (J) HC_Abuse_Dependence_Psychosis | Mean Difference (I-J) | Std. Error | Sig.[b] | 95% Confidence Interval for Difference[b] Lower Bound | Upper Bound |
|---|---|---|---|---|---|---|
| 0 | 1 | 0.127 | 0.229 | 0.582 | -0.326 | 0.579 |
| | 2 | 0.246 | 0.227 | 0.281 | -0.203 | 0.694 |
| | 3 | .823* | 0.226 | 0.000 | 0.377 | 1.269 |
| 1 | 0 | -0.127 | 0.229 | 0.582 | -0.579 | 0.326 |
| | 2 | 0.119 | 0.198 | 0.549 | -0.272 | 0.509 |
| | 3 | .696* | 0.210 | 0.001 | 0.282 | 1.110 |
| 2 | 0 | -0.246 | 0.227 | 0.281 | -0.694 | 0.203 |
| | 1 | -0.119 | 0.198 | 0.549 | -0.509 | 0.272 |
| | 3 | .577* | 0.205 | 0.006 | 0.172 | 0.983 |
| 3 | 0 | -.823* | 0.226 | 0.000 | -1.269 | -0.377 |
| | 1 | -.696* | 0.210 | 0.001 | -1.110 | -0.282 |
| | 2 | -.577* | 0.205 | 0.006 | -0.983 | -0.172 |

Based on estimated marginal means

*. The mean difference is significant at the 0.05 level.

b. Adjustment for multiple comparisons: Least Significant Difference (equivalent to no adjustments).

HC=0, MA use=1, MA dependence=2, MAP=3

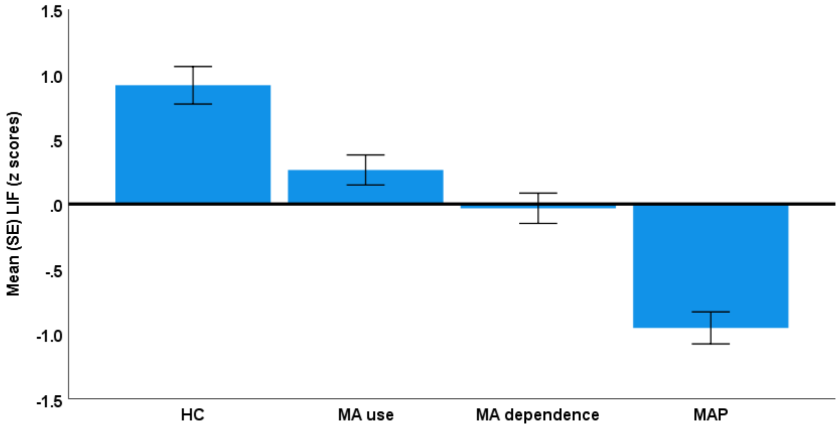


Figure S21. LIF levels in healthy controls (HC) and people with methamphetamine (MA) use, MA dependence, and MA-induced psychosis (MAP) (F=35.84, df=3/164, p<0.001).

**Pairwise Comparisons**

Dependent Variable: LIF

| (I) HC_Abuse_ Dependence_ Psychosis | (J) HC_Abuse_ Dependence_ Psychosis | Mean Difference (I-J) | Std. Error | Sig.[b] | 95% Confidence Interval for Difference[b] | |
|---|---|---|---|---|---|---|
| | | | | | Lower Bound | Upper Bound |
| 0 | 1 | .652* | 0.190 | 0.001 | 0.276 | 1.028 |
| | 2 | .947* | 0.189 | 0.000 | 0.575 | 1.320 |
| | 3 | 1.869* | 0.187 | 0.000 | 1.499 | 2.239 |
| 1 | 0 | -.652* | 0.190 | 0.001 | -1.028 | -0.276 |
| | 2 | 0.295 | 0.164 | 0.074 | -0.029 | 0.619 |
| | 3 | 1.217* | 0.174 | 0.000 | 0.873 | 1.560 |
| 2 | 0 | -.947* | 0.189 | 0.000 | -1.320 | -0.575 |
| | 1 | -0.295 | 0.164 | 0.074 | -0.619 | 0.029 |
| | 3 | .921* | 0.171 | 0.000 | 0.585 | 1.258 |
| 3 | 0 | -1.869* | 0.187 | 0.000 | -2.239 | -1.499 |
| | 1 | -1.217* | 0.174 | 0.000 | -1.560 | -0.873 |
| | 2 | -.921* | 0.171 | 0.000 | -1.258 | -0.585 |

Based on estimated marginal means

*. The mean difference is significant at the 0.05 level.

b. Adjustment for multiple comparisons: Least Significant Difference (equivalent to no adjustments).

HC=0, MA use=1, MA dependence=2, MAP=3

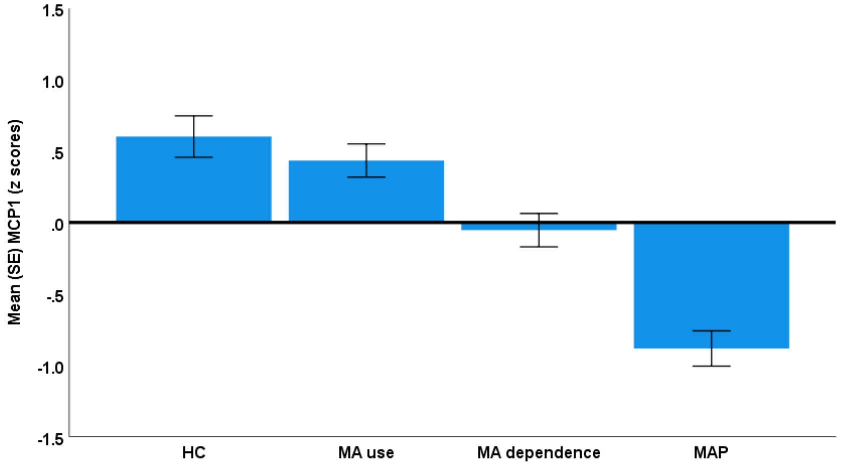


Figure S22. CCL2 (MCP1) levels in healthy controls (HC) and people with methamphetamine (MA) use, MA dependence, and MA-induced psychosis (MAP) (F=27.62, df=3/164, p<0.001).

**Pairwise Comparisons**

Dependent Variable: MCP1

| (I) HC_Abuse_Dependence_Psychosis | (J) HC_Abuse_Dependence_Psychosis | Mean Difference (I-J) | Std. Error | Sig.[b] | 95% Confidence Interval for Difference[b] | |
|---|---|---|---|---|---|---|
| | | | | | Lower Bound | Upper Bound |
| 0 | 1 | 0.168 | 0.191 | 0.381 | -0.209 | 0.545 |
| | 2 | .656* | 0.189 | 0.001 | 0.282 | 1.029 |
| | 3 | 1.485* | 0.188 | 0.000 | 1.114 | 1.856 |
| 1 | 0 | -0.168 | 0.191 | 0.381 | -0.545 | 0.209 |
| | 2 | .488* | 0.165 | 0.003 | 0.163 | 0.813 |
| | 3 | 1.317* | 0.175 | 0.000 | 0.972 | 1.662 |
| 2 | 0 | -.656* | 0.189 | 0.001 | -1.029 | -0.282 |
| | 1 | -.488* | 0.165 | 0.003 | -0.813 | -0.163 |
| | 3 | .829* | 0.171 | 0.000 | 0.491 | 1.167 |
| 3 | 0 | -1.485* | 0.188 | 0.000 | -1.856 | -1.114 |
| | 1 | -1.317* | 0.175 | 0.000 | -1.662 | -0.972 |
| | 2 | -.829* | 0.171 | 0.000 | -1.167 | -0.491 |

Based on estimated marginal means

*. The mean difference is significant at the 0.05 level.

b. Adjustment for multiple comparisons: Least Significant Difference (equivalent to no adjustments).

HC=0, MA use=1, MA dependence=2, MAP=3

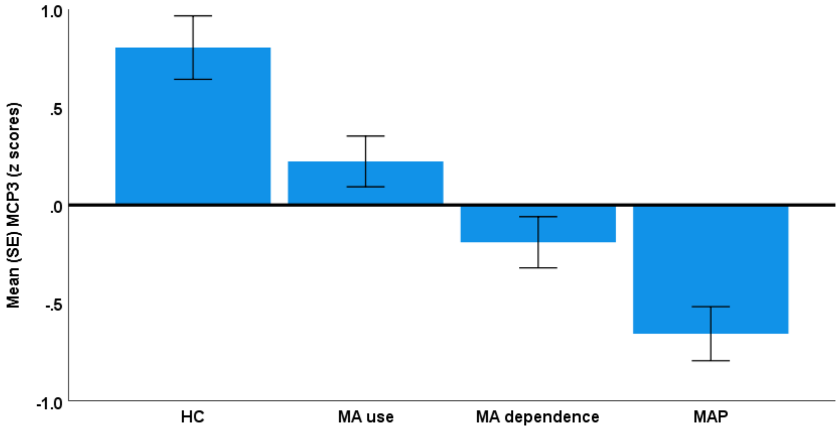


Figure S23. CCL7 (MCP3) levels in healthy controls (HC) and people with methamphetamine (MA) use, MA dependence, and MA-induced psychosis (MAP) (F=17.83, df=3/164, p<0.001).

**Pairwise Comparisons**

Dependent Variable: MCP3

| (I) HC_Abuse_Dependence_Psychosis | (J) HC_Abuse_Dependence_Psychosis | Mean Difference (I-J) | Std. Error | Sig.[b] | 95% Confidence Interval for Difference[b] | |
|---|---|---|---|---|---|---|
| | | | | | Lower Bound | Upper Bound |
| 0 | 1 | .582* | 0.213 | 0.007 | 0.160 | 1.003 |
| | 2 | .995* | 0.211 | 0.000 | 0.578 | 1.412 |
| | 3 | 1.462* | 0.210 | 0.000 | 1.047 | 1.876 |
| 1 | 0 | -.582* | 0.213 | 0.007 | -1.003 | -0.160 |
| | 2 | .413* | 0.184 | 0.026 | 0.050 | 0.776 |
| | 3 | .880* | 0.195 | 0.000 | 0.495 | 1.265 |
| 2 | 0 | -.995* | 0.211 | 0.000 | -1.412 | -0.578 |
| | 1 | -.413* | 0.184 | 0.026 | -0.776 | -0.050 |
| | 3 | .467* | 0.191 | 0.016 | 0.090 | 0.844 |
| 3 | 0 | -1.462* | 0.210 | 0.000 | -1.876 | -1.047 |
| | 1 | -.880* | 0.195 | 0.000 | -1.265 | -0.495 |
| | 2 | -.467* | 0.191 | 0.016 | -0.844 | -0.090 |

Based on estimated marginal means

*. The mean difference is significant at the 0.05 level.

b. Adjustment for multiple comparisons: Least Significant Difference (equivalent to no adjustments).

HC=0, MA use=1, MA dependence=2, MAP=3

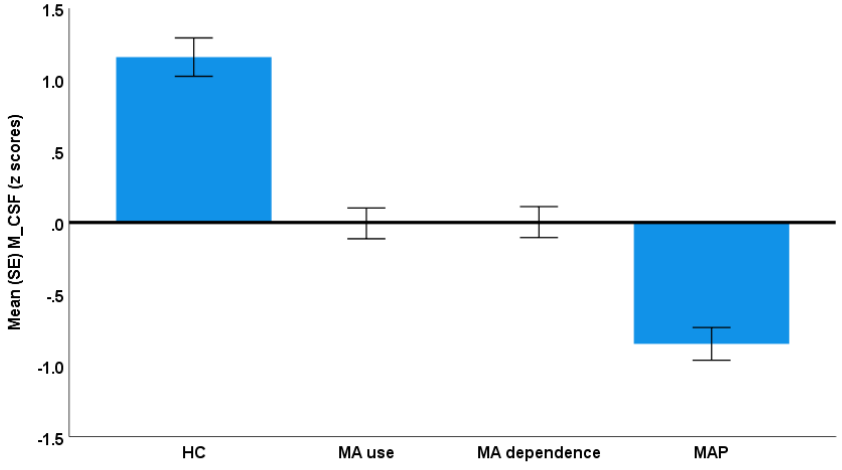


Figure S24. CSF levels in healthy controls (HC) and people with methamphetamine (MA) use, MA dependence, and MA-induced psychosis (MAP) (F=44.36, df=3/164, p<0.001).

**Pairwise Comparisons**

Dependent Variable: CSF

| (I) HC_Abuse_ Dependence_ Psychosis | (J) HC_Abuse_ Dependence_ Psychosis | Mean Difference (I-J) | Std. Error | Sig.[b] | 95% Confidence Interval for Difference[b] | |
|---|---|---|---|---|---|---|
| | | | | | Lower Bound | Upper Bound |
| 0 | 1 | 1.164* | 0.177 | 0.000 | 0.815 | 1.514 |
| | 2 | 1.156* | 0.175 | 0.000 | 0.810 | 1.502 |
| | 3 | 2.009* | 0.174 | 0.000 | 1.665 | 2.353 |
| 1 | 0 | -1.164* | 0.177 | 0.000 | -1.514 | -0.815 |
| | 2 | -0.009 | 0.153 | 0.954 | -0.310 | 0.293 |
| | 3 | .844* | 0.162 | 0.000 | 0.525 | 1.164 |
| 2 | 0 | -1.156* | 0.175 | 0.000 | -1.502 | -0.810 |
| | 1 | 0.009 | 0.153 | 0.954 | -0.293 | 0.310 |
| | 3 | .853* | 0.159 | 0.000 | 0.540 | 1.166 |
| 3 | 0 | -2.009* | 0.174 | 0.000 | -2.353 | -1.665 |
| | 1 | -.844* | 0.162 | 0.000 | -1.164 | -0.525 |
| | 2 | -.853* | 0.159 | 0.000 | -1.166 | -0.540 |

Based on estimated marginal means

*. The mean difference is significant at the 0.05 level.

b. Adjustment for multiple comparisons: Least Significant Difference (equivalent to no adjustments).

HC=0, MA use=1, MA dependence=2, MAP=3

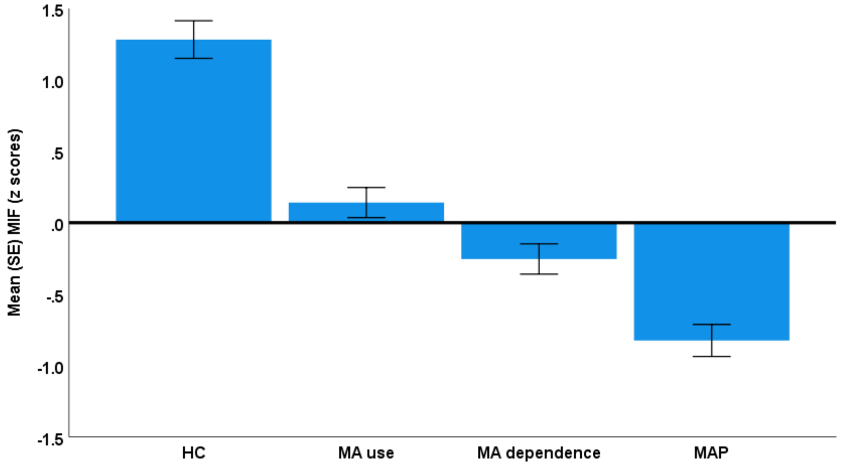


Figure S25. MIF levels in healthy controls (HC) and people with methamphetamine (MA) use, MA dependence, and MA-induced psychosis (MAP) (F=53.30, df=3/164, p<0.001).

**Pairwise Comparisons**

Dependent Variable: MIF

| (I) HC_Abuse_ Dependence_ Psychosis | (J) HC_Abuse_ Dependence_ Psychosis | Mean Difference (I-J) | Std. Error | Sig.[b] | 95% Confidence Interval for Difference[b] | |
|---|---|---|---|---|---|---|
| | | | | | Lower Bound | Upper Bound |
| 0 | 1 | 1.141* | 0.173 | 0.000 | 0.799 | 1.484 |
| | 2 | 1.537* | 0.172 | 0.000 | 1.198 | 1.876 |
| | 3 | 2.107* | 0.171 | 0.000 | 1.771 | 2.444 |
| 1 | 0 | -1.141* | 0.173 | 0.000 | -1.484 | -0.799 |
| | 2 | .396* | 0.149 | 0.009 | 0.100 | 0.691 |
| | 3 | .966* | 0.159 | 0.000 | 0.653 | 1.279 |
| 2 | 0 | -1.537* | 0.172 | 0.000 | -1.876 | -1.198 |
| | 1 | -.396* | 0.149 | 0.009 | -0.691 | -0.100 |
| | 3 | .570* | 0.155 | 0.000 | 0.264 | 0.877 |
| 3 | 0 | -2.107* | 0.171 | 0.000 | -2.444 | -1.771 |
| | 1 | -.966* | 0.159 | 0.000 | -1.279 | -0.653 |
| | 2 | -.570* | 0.155 | 0.000 | -0.877 | -0.264 |

Based on estimated marginal means

*. The mean difference is significant at the 0.05 level.

b. Adjustment for multiple comparisons: Least Significant Difference (equivalent to no adjustments).

HC=0, MA use=1, MA dependence=2, MAP=3

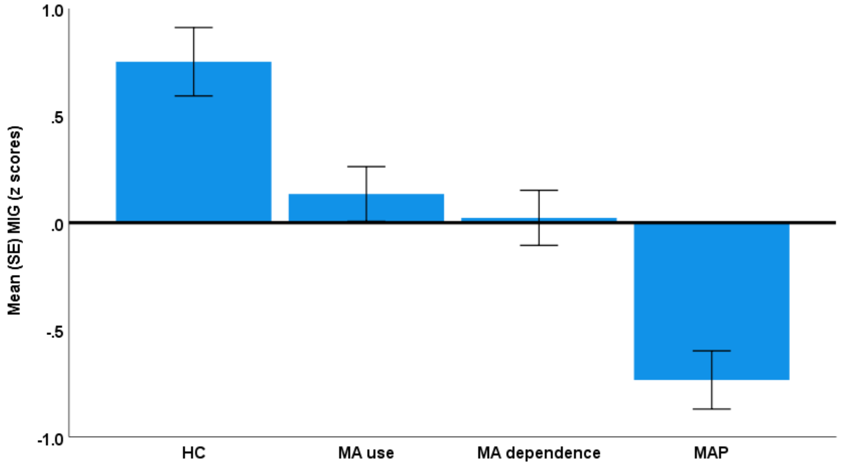


Figure S26. CXCL9 or MIG levels in healthy controls (HC) and people with methamphetamine (MA) use, MA dependence, and MA-induced psychosis (MAP) (F=17.96, df=3/164, p<0.001).

**Pairwise Comparisons**

Dependent Variable: MIG

| (I) HC_Abuse_ Dependence_ Psychosis | (J) HC_Abuse_ Dependence_ Psychosis | Mean Difference (I-J) | Std. Error | Sig.[b] | 95% Confidence Interval for Difference[b] | |
|---|---|---|---|---|---|---|
| | | | | | Lower Bound | Upper Bound |
| 0 | 1 | .618* | 0.210 | 0.004 | 0.203 | 1.033 |
| | 2 | .729* | 0.208 | 0.001 | 0.318 | 1.140 |
| | 3 | 1.486* | 0.207 | 0.000 | 1.078 | 1.894 |
| 1 | 0 | -.618* | 0.210 | 0.004 | -1.033 | -0.203 |
| | 2 | 0.111 | 0.181 | 0.540 | -0.246 | 0.469 |
| | 3 | .868* | 0.192 | 0.000 | 0.489 | 1.247 |
| 2 | 0 | -.729* | 0.208 | 0.001 | -1.140 | -0.318 |
| | 1 | -0.111 | 0.181 | 0.540 | -0.469 | 0.246 |
| | 3 | .757* | 0.188 | 0.000 | 0.385 | 1.128 |
| 3 | 0 | -1.486* | 0.207 | 0.000 | -1.894 | -1.078 |
| | 1 | -.868* | 0.192 | 0.000 | -1.247 | -0.489 |
| | 2 | -.757* | 0.188 | 0.000 | -1.128 | -0.385 |

Based on estimated marginal means

*. The mean difference is significant at the 0.05 level.

b. Adjustment for multiple comparisons: Least Significant Difference (equivalent to no adjustments).

HC=0, MA use=1, MA dependence=2, MAP=3

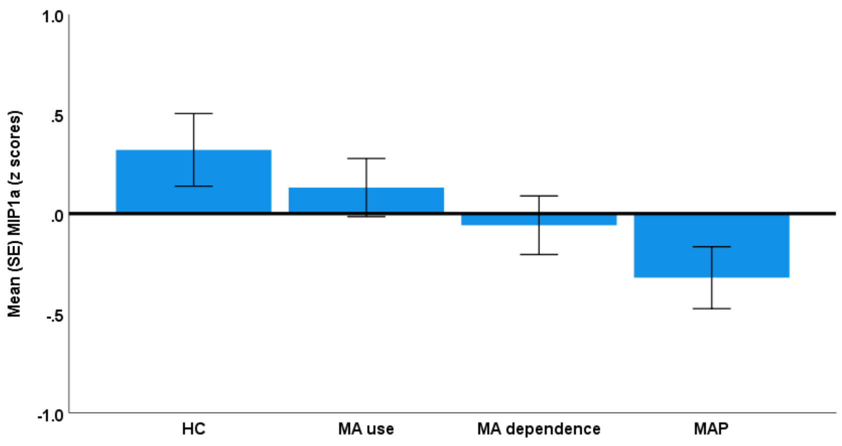


Figure S27. CCL3 or (MIP-1α) levels in healthy controls (HC) and people with methamphetamine (MA) use, MA dependence, and MA-induced psychosis (MAP) (F=2.83, df=3/164, p=0.040).

**Pairwise Comparisons**

Dependent Variable: MIP-1α

| (I) HC_Abuse_ Dependence_ Psychosis | (J) HC_Abuse_ Dependence_ Psychosis | Mean Difference (I-J) | Std. Error | Sig.[b] | 95% Confidence Interval for Difference[b] | |
|---|---|---|---|---|---|---|
| | | | | | Lower Bound | Upper Bound |
| 0 | 1 | 0.189 | 0.240 | 0.431 | -0.285 | 0.663 |
| | 2 | 0.378 | 0.238 | 0.113 | -0.091 | 0.848 |
| | 3 | .642* | 0.236 | 0.007 | 0.175 | 1.108 |
| 1 | 0 | -0.189 | 0.240 | 0.431 | -0.663 | 0.285 |
| | 2 | 0.189 | 0.207 | 0.362 | -0.220 | 0.598 |
| | 3 | .452* | 0.219 | 0.041 | 0.019 | 0.886 |
| 2 | 0 | -0.378 | 0.238 | 0.113 | -0.848 | 0.091 |
| | 1 | -0.189 | 0.207 | 0.362 | -0.598 | 0.220 |
| | 3 | 0.263 | 0.215 | 0.222 | -0.161 | 0.688 |
| 3 | 0 | -.642* | 0.236 | 0.007 | -1.108 | -0.175 |
| | 1 | -.452* | 0.219 | 0.041 | -0.886 | -0.019 |
| | 2 | -0.263 | 0.215 | 0.222 | -0.688 | 0.161 |

Based on estimated marginal means

*. The mean difference is significant at the 0.05 level.

b. Adjustment for multiple comparisons: Least Significant Difference (equivalent to no adjustments).

HC=0, MA use=1, MA dependence=2, MAP=3

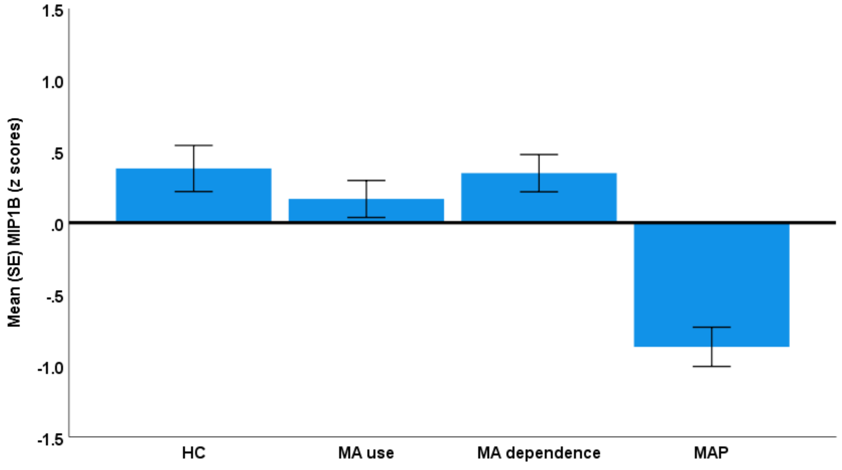


Figure S28. CCL4 or MIP-1β levels in healthy controls (HC) and people with methamphetamine (MA) use, MA dependence, and MA-induced psychosis (MAP) (F=18.20, df=3/164, p<0.001).

**Pairwise Comparisons**

Dependent Variable: MIP-1β

| (I) HC_Abuse_ Dependence_ Psychosis | (J) HC_Abuse_ Dependence_ Psychosis | Mean Difference (I-J) | Std. Error | Sig.[b] | 95% Confidence Interval for Difference[b] | |
|---|---|---|---|---|---|---|
| | | | | | Lower Bound | Upper Bound |
| 0 | 1 | 0.213 | 0.213 | 0.317 | -0.206 | 0.633 |
| | 2 | 0.033 | 0.211 | 0.877 | -0.383 | 0.448 |
| | 3 | 1.249* | 0.209 | 0.000 | 0.836 | 1.662 |
| 1 | 0 | -0.213 | 0.213 | 0.317 | -0.633 | 0.206 |
| | 2 | -0.181 | 0.183 | 0.326 | -0.543 | 0.181 |
| | 3 | 1.036* | 0.194 | 0.000 | 0.652 | 1.419 |
| 2 | 0 | -0.033 | 0.211 | 0.877 | -0.448 | 0.383 |
| | 1 | 0.181 | 0.183 | 0.326 | -0.181 | 0.543 |
| | 3 | 1.216* | 0.190 | 0.000 | 0.840 | 1.592 |
| 3 | 0 | -1.249* | 0.209 | 0.000 | -1.662 | -0.836 |
| | 1 | -1.036* | 0.194 | 0.000 | -1.419 | -0.652 |
| | 2 | -1.216* | 0.190 | 0.000 | -1.592 | -0.840 |

Based on estimated marginal means

*. The mean difference is significant at the 0.05 level.

b. Adjustment for multiple comparisons: Least Significant Difference (equivalent to no adjustments).

HC=0, MA use=1, MA dependence=2, MAP=3

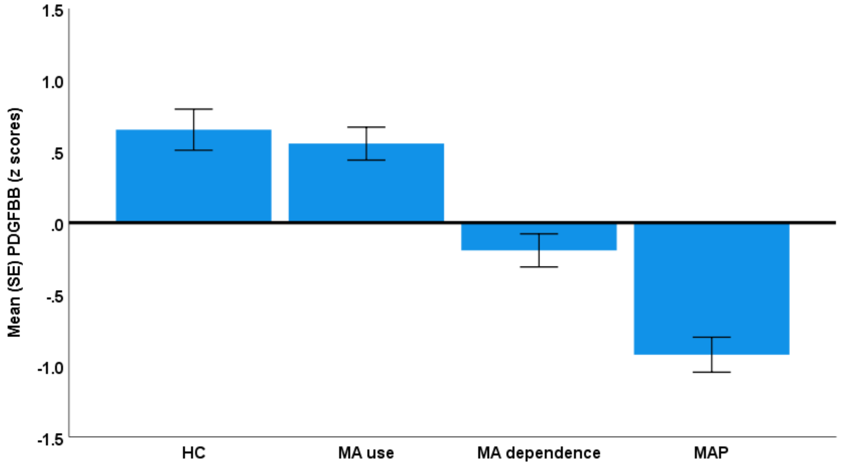


Figure S29. PDGF-BB levels in healthy controls (HC) and people with methamphetamine (MA) use, MA dependence, and MA-induced psychosis (MAP) (F=34.46, df=3/164, p<0.001).

**Pairwise Comparisons**

Dependent Variable: PDGFBB

| (I) HC_Abuse_Dependence_Psychosis | (J) HC_Abuse_Dependence_Psychosis | Mean Difference (I-J) | Std. Error | Sig.[b] | 95% Confidence Interval for Difference[b] | |
|---|---|---|---|---|---|---|
| | | | | | Lower Bound | Upper Bound |
| 0 | 1 | 0.097 | 0.189 | 0.608 | -0.277 | 0.472 |
| | 2 | .845* | 0.188 | 0.000 | 0.475 | 1.216 |
| | 3 | 1.576* | 0.186 | 0.000 | 1.208 | 1.944 |
| 1 | 0 | -0.097 | 0.189 | 0.608 | -0.472 | 0.277 |
| | 2 | .748* | 0.163 | 0.000 | 0.425 | 1.070 |
| | 3 | 1.479* | 0.173 | 0.000 | 1.137 | 1.821 |
| 2 | 0 | -.845* | 0.188 | 0.000 | -1.216 | -0.475 |
| | 1 | -.748* | 0.163 | 0.000 | -1.070 | -0.425 |
| | 3 | .731* | 0.170 | 0.000 | 0.396 | 1.066 |
| 3 | 0 | -1.576* | 0.186 | 0.000 | -1.944 | -1.208 |
| | 1 | -1.479* | 0.173 | 0.000 | -1.821 | -1.137 |
| | 2 | -.731* | 0.170 | 0.000 | -1.066 | -0.396 |

Based on estimated marginal means

*. The mean difference is significant at the 0.05 level.

b. Adjustment for multiple comparisons: Least Significant Difference (equivalent to no adjustments).

HC=0, MA use=1, MA dependence=2, MAP=3

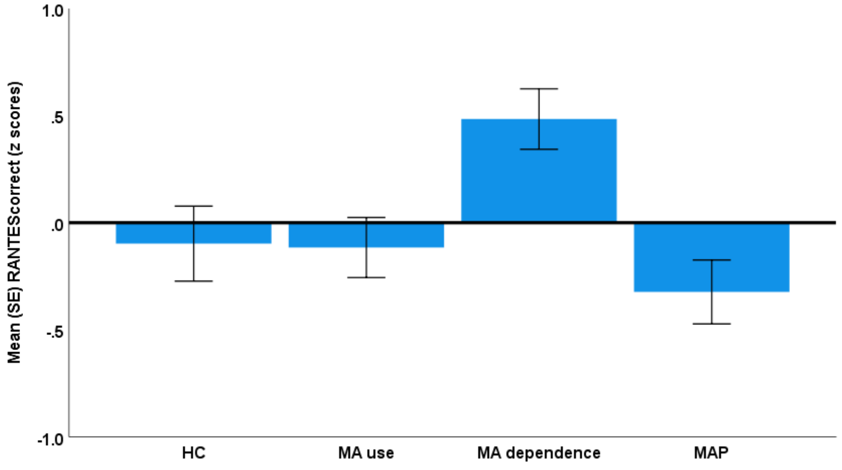


Figure S30. CCL5 or RANTES levels in healthy controls (HC) and people with methamphetamine (MA) use, MA dependence, and MA-induced psychosis (MAP) (F=5.81, df=3/164, p<0.001).

**Pairwise Comparisons**

Dependent Variable: RANTES

| (I) HC_Abuse_ Dependence_ Psychosis | (J) HC_Abuse_ Dependence_ Psychosis | Mean Difference (I-J) | Std. Error | Sig.[b] | 95% Confidence Interval for Difference[b] | |
|---|---|---|---|---|---|---|
| | | | | | Lower Bound | Upper Bound |
| 0 | 1 | 0.018 | 0.231 | 0.937 | -0.437 | 0.473 |
| | 2 | -.582* | 0.228 | 0.012 | -1.033 | -0.131 |
| | 3 | 0.226 | 0.227 | 0.321 | -0.222 | 0.674 |
| 1 | 0 | -0.018 | 0.231 | 0.937 | -0.473 | 0.437 |
| | 2 | -.600* | 0.199 | 0.003 | -0.993 | -0.207 |
| | 3 | 0.207 | 0.211 | 0.327 | -0.209 | 0.624 |
| 2 | 0 | .582* | 0.228 | 0.012 | 0.131 | 1.033 |
| | 1 | .600* | 0.199 | 0.003 | 0.207 | 0.993 |
| | 3 | .807* | 0.207 | 0.000 | 0.400 | 1.215 |
| 3 | 0 | -0.226 | 0.227 | 0.321 | -0.674 | 0.222 |
| | 1 | -0.207 | 0.211 | 0.327 | -0.624 | 0.209 |
| | 2 | -.807* | 0.207 | 0.000 | -1.215 | -0.400 |

Based on estimated marginal means

*. The mean difference is significant at the 0.05 level.

b. Adjustment for multiple comparisons: Least Significant Difference (equivalent to no adjustments).

HC=0, MA use=1, MA dependence=2, MAP=3

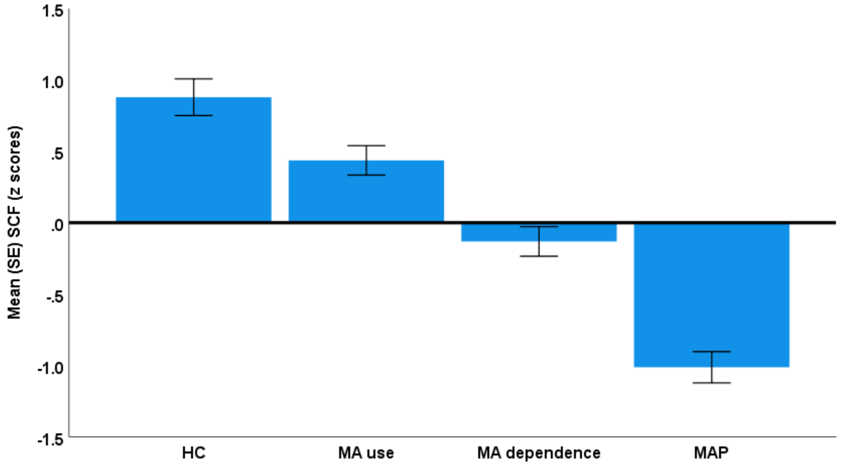


Figure S31. SCF levels in healthy controls (HC) and people with methamphetamine (MA) use, MA dependence, and MA-induced psychosis (MAP) ($F=51.39$, $df=3/164$, $p<0.001$).

**Pairwise Comparisons**

Dependent Variable: SCF

| (I) HC_Abuse_ Dependence_ Psychosis | (J) HC_Abuse_ Dependence_ Psychosis | Mean Difference (I-J) | Std. Error | Sig.[b] | 95% Confidence Interval for Difference[b] | |
|---|---|---|---|---|---|---|
| | | | | | Lower Bound | Upper Bound |
| 0 | 1 | .443* | 0.169 | 0.010 | 0.109 | 0.776 |
| | 2 | 1.010* | 0.167 | 0.000 | 0.679 | 1.341 |
| | 3 | 1.892* | 0.166 | 0.000 | 1.563 | 2.220 |
| 1 | 0 | -.443* | 0.169 | 0.010 | -0.776 | -0.109 |
| | 2 | .567* | 0.146 | 0.000 | 0.280 | 0.855 |
| | 3 | 1.449* | 0.155 | 0.000 | 1.144 | 1.754 |
| 2 | 0 | -1.010* | 0.167 | 0.000 | -1.341 | -0.679 |
| | 1 | -.567* | 0.146 | 0.000 | -0.855 | -0.280 |
| | 3 | .882* | 0.151 | 0.000 | 0.582 | 1.181 |
| 3 | 0 | -1.892* | 0.166 | 0.000 | -2.220 | -1.563 |
| | 1 | -1.449* | 0.155 | 0.000 | -1.754 | -1.144 |
| | 2 | -.882* | 0.151 | 0.000 | -1.181 | -0.582 |

Based on estimated marginal means

*. The mean difference is significant at the 0.05 level.

b. Adjustment for multiple comparisons: Least Significant Difference (equivalent to no adjustments).

HC=0, MA use=1, MA dependence=2, MAP=3

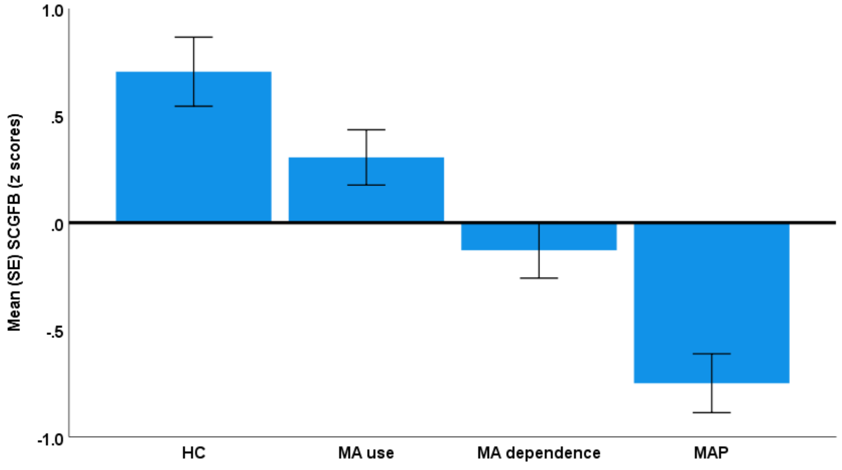


Figure S32. SCGF-β levels in healthy controls (HC) and people with methamphetamine (MA) use, MA dependence, and MA-induced psychosis (MAP) (F=18.79, df=3/164, p<0.001).

**Pairwise Comparisons**

Dependent Variable: SCGF-β

| (I) HC_Abuse_ Dependence_ Psychosis | (J) HC_Abuse_ Dependence_ Psychosis | Mean Difference (I-J) | Std. Error | Sig.[b] | 95% Confidence Interval for Difference[b] | |
|---|---|---|---|---|---|---|
| | | | | | Lower Bound | Upper Bound |
| 0 | 1 | 0.400 | 0.212 | 0.061 | -0.019 | 0.819 |
| | 2 | .834* | 0.210 | 0.000 | 0.419 | 1.249 |
| | 3 | 1.454* | 0.209 | 0.000 | 1.042 | 1.867 |
| 1 | 0 | -0.400 | 0.212 | 0.061 | -0.819 | 0.019 |
| | 2 | .434* | 0.183 | 0.019 | 0.072 | 0.795 |
| | 3 | 1.054* | 0.194 | 0.000 | 0.671 | 1.437 |
| 2 | 0 | -.834* | 0.210 | 0.000 | -1.249 | -0.419 |
| | 1 | -.434* | 0.183 | 0.019 | -0.795 | -0.072 |
| | 3 | .621* | 0.190 | 0.001 | 0.245 | 0.996 |
| 3 | 0 | -1.454* | 0.209 | 0.000 | -1.867 | -1.042 |
| | 1 | -1.054* | 0.194 | 0.000 | -1.437 | -0.671 |
| | 2 | -.621* | 0.190 | 0.001 | -0.996 | -0.245 |

Based on estimated marginal means

*. The mean difference is significant at the 0.05 level.

b. Adjustment for multiple comparisons: Least Significant Difference (equivalent to no adjustments).

HC=0, MA use=1, MA dependence=2, MAP=3

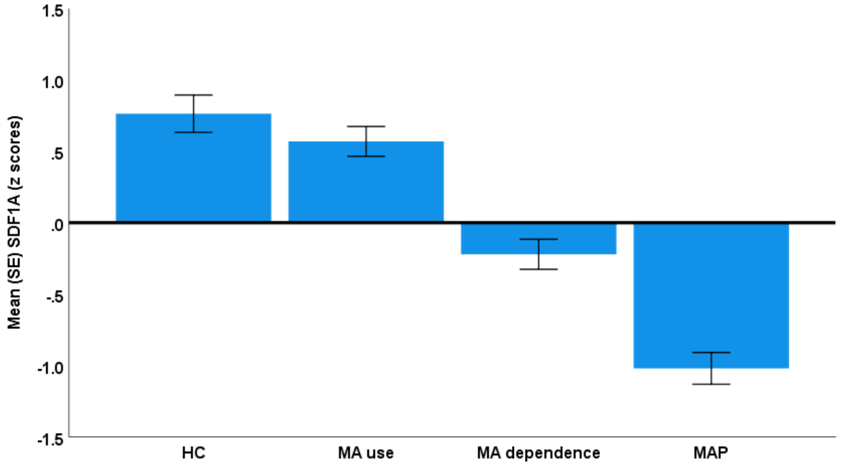


Figure S33. CXCL12 or SDF-1α levels in healthy controls (HC) and people with methamphetamine (MA) use, MA dependence, and MA-induced psychosis (MAP) (F=50.94, df=3/164, p<0.001).

**Pairwise Comparisons**

Dependent Variable: SDF-1α

| (I) HC_Abuse_ Dependence_ Psychosis | (J) HC_Abuse_ Dependence_ Psychosis | Mean Difference (I-J) | Std. Error | Sig.[b] | 95% Confidence Interval for Difference[b] | |
|---|---|---|---|---|---|---|
| | | | | | Lower Bound | Upper Bound |
| 0 | 1 | 0.194 | 0.172 | 0.260 | -0.145 | 0.534 |
| | 2 | .985* | 0.170 | 0.000 | 0.648 | 1.321 |
| | 3 | 1.784* | 0.169 | 0.000 | 1.450 | 2.118 |
| 1 | 0 | -0.194 | 0.172 | 0.260 | -0.534 | 0.145 |
| | 2 | .790* | 0.148 | 0.000 | 0.497 | 1.083 |
| | 3 | 1.590* | 0.157 | 0.000 | 1.279 | 1.900 |
| 2 | 0 | -.985* | 0.170 | 0.000 | -1.321 | -0.648 |
| | 1 | -.790* | 0.148 | 0.000 | -1.083 | -0.497 |
| | 3 | .799* | 0.154 | 0.000 | 0.495 | 1.103 |
| 3 | 0 | -1.784* | 0.169 | 0.000 | -2.118 | -1.450 |
| | 1 | -1.590* | 0.157 | 0.000 | -1.900 | -1.279 |
| | 2 | -.799* | 0.154 | 0.000 | -1.103 | -0.495 |

Based on estimated marginal means

*. The mean difference is significant at the 0.05 level.

b. Adjustment for multiple comparisons: Least Significant Difference (equivalent to no adjustments).

HC=0, MA use=1, MA dependence=2, MAP=3

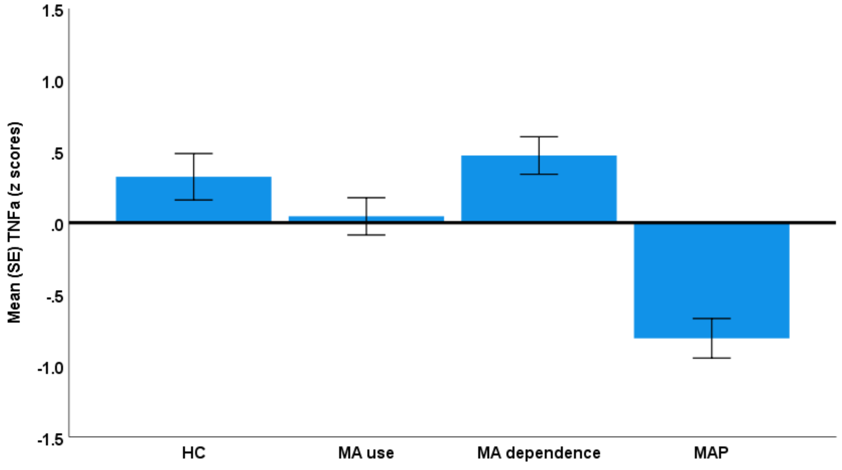


Figure S34. TNF-α levels in healthy controls (HC) and people with methamphetamine (MA) use, MA dependence, and MA-induced psychosis (MAP) (F=17.20, df=3/164, p<0.001).

**Pairwise Comparisons**

Dependent Variable: TNF-α

| (I) HC_Abuse_ Dependence_ Psychosis | (J) HC_Abuse_ Dependence_ Psychosis | Mean Difference (I-J) | Std. Error | Sig.[b] | 95% Confidence Interval for Difference[b] | |
|---|---|---|---|---|---|---|
| | | | | | Lower Bound | Upper Bound |
| 0 | 1 | 0.277 | 0.214 | 0.197 | -0.146 | 0.700 |
| | 2 | -0.149 | 0.212 | 0.484 | -0.568 | 0.270 |
| | 3 | 1.131* | 0.211 | 0.000 | 0.715 | 1.548 |
| 1 | 0 | -0.277 | 0.214 | 0.197 | -0.700 | 0.146 |
| | 2 | -.426* | 0.185 | 0.022 | -0.791 | -0.061 |
| | 3 | .854* | 0.196 | 0.000 | 0.467 | 1.241 |
| 2 | 0 | 0.149 | 0.212 | 0.484 | -0.270 | 0.568 |
| | 1 | .426* | 0.185 | 0.022 | 0.061 | 0.791 |
| | 3 | 1.280* | 0.192 | 0.000 | 0.901 | 1.659 |
| 3 | 0 | -1.131* | 0.211 | 0.000 | -1.548 | -0.715 |
| | 1 | -.854* | 0.196 | 0.000 | -1.241 | -0.467 |
| | 2 | -1.280* | 0.192 | 0.000 | -1.659 | -0.901 |

Based on estimated marginal means

*. The mean difference is significant at the 0.05 level.

b. Adjustment for multiple comparisons: Least Significant Difference (equivalent to no adjustments).

HC=0, MA use=1, MA dependence=2, MAP=3

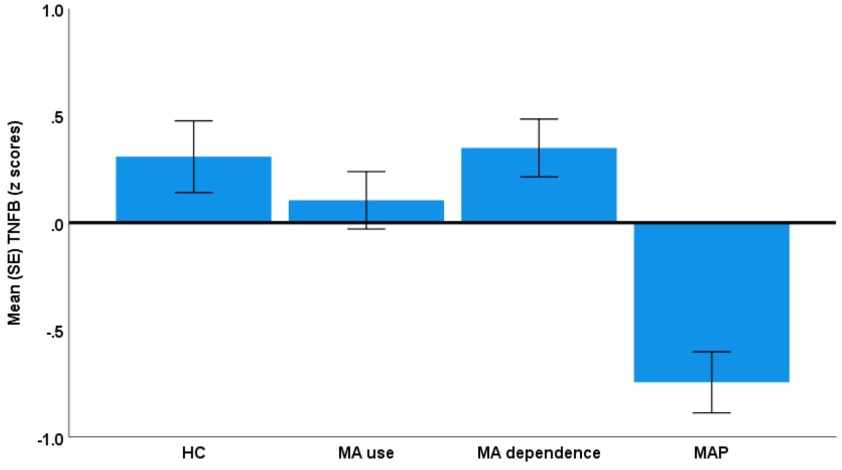


Figure S35. TNF-β levels in healthy controls (HC) and people with methamphetamine (MA) use, MA dependence, and MA-induced psychosis (MAP) ($F=12.78$, $df=3/164$, $p<0.001$).

**Pairwise Comparisons**

Dependent Variable: TNF-β

| (I) HC_Abuse_Dependence_Psychosis | (J) HC_Abuse_Dependence_Psychosis | Mean Difference (I-J) | Std. Error | Sig.[b] | 95% Confidence Interval for Difference[b] | |
|---|---|---|---|---|---|---|
| | | | | | Lower Bound | Upper Bound |
| 0 | 1 | 0.204 | 0.220 | 0.357 | -0.231 | 0.639 |
| | 2 | -0.041 | 0.218 | 0.853 | -0.471 | 0.390 |
| | 3 | 1.053* | 0.217 | 0.000 | 0.626 | 1.481 |
| 1 | 0 | -0.204 | 0.220 | 0.357 | -0.639 | 0.231 |
| | 2 | -0.244 | 0.190 | 0.200 | -0.619 | 0.131 |
| | 3 | .850* | 0.201 | 0.000 | 0.452 | 1.248 |
| 2 | 0 | 0.041 | 0.218 | 0.853 | -0.390 | 0.471 |
| | 1 | 0.244 | 0.190 | 0.200 | -0.131 | 0.619 |
| | 3 | 1.094* | 0.197 | 0.000 | 0.704 | 1.484 |
| 3 | 0 | -1.053* | 0.217 | 0.000 | -1.481 | -0.626 |
| | 1 | -.850* | 0.201 | 0.000 | -1.248 | -0.452 |
| | 2 | -1.094* | 0.197 | 0.000 | -1.484 | -0.704 |

Based on estimated marginal means

*. The mean difference is significant at the 0.05 level.

b. Adjustment for multiple comparisons: Least Significant Difference (equivalent to no adjustments).

HC=0, MA use=1, MA dependence=2, MAP=3

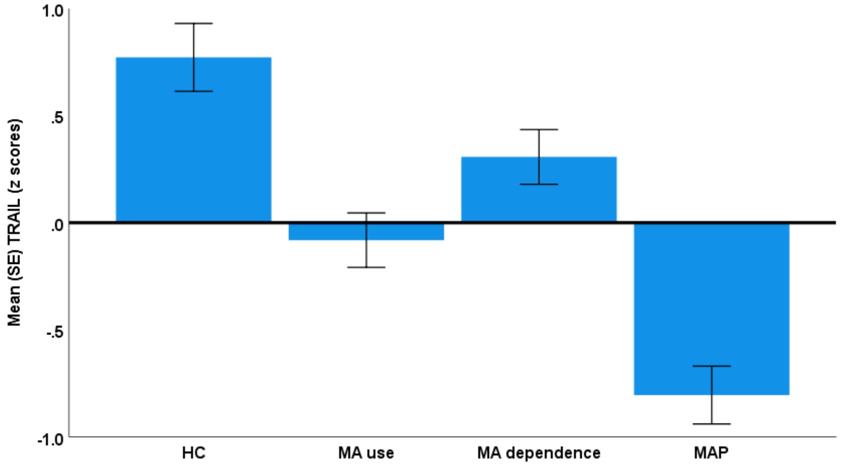


Figure S36. TRAIL levels in healthy controls (HC) and people with methamphetamine (MA) use, MA dependence, and MA-induced psychosis (MAP) (F=22.53, df=3/164, p<0.001).

**Pairwise Comparisons**

Dependent Variable: TRAIL

| (I) HC_Abuse_ Dependence_ Psychosis | (J) HC_Abuse_ Dependence_ Psychosis | Mean Difference (I-J) | Std. Error | Sig.[b] | 95% Confidence Interval for Difference[b] | |
|---|---|---|---|---|---|---|
| | | | | | Lower Bound | Upper Bound |
| 0 | 1 | .854* | 0.209 | 0.000 | 0.441 | 1.266 |
| | 2 | .465* | 0.207 | 0.026 | 0.057 | 0.873 |
| | 3 | 1.577* | 0.205 | 0.000 | 1.171 | 1.982 |
| 1 | 0 | -.854* | 0.209 | 0.000 | -1.266 | -0.441 |
| | 2 | -.389* | 0.180 | 0.032 | -0.744 | -0.033 |
| | 3 | .723* | 0.191 | 0.000 | 0.346 | 1.100 |
| 2 | 0 | -.465* | 0.207 | 0.026 | -0.873 | -0.057 |
| | 1 | .389* | 0.180 | 0.032 | 0.033 | 0.744 |
| | 3 | 1.112* | 0.187 | 0.000 | 0.743 | 1.481 |
| 3 | 0 | -1.577* | 0.205 | 0.000 | -1.982 | -1.171 |
| | 1 | -.723* | 0.191 | 0.000 | -1.100 | -0.346 |
| | 2 | -1.112* | 0.187 | 0.000 | -1.481 | -0.743 |

Based on estimated marginal means

*. The mean difference is significant at the 0.05 level.

b. Adjustment for multiple comparisons: Least Significant Difference (equivalent to no adjustments).

HC=0, MA use=1, MA dependence=2, MAP=3
